# Trends and inequalities in Advice and Guidance versus direct referral in NHS primary care: a population-based study, 2015–2023

**DOI:** 10.1101/2025.09.25.25336644

**Authors:** Kayleigh J Mason, Kelvin P Jordan, James Bailey, Ram Bajpai, Lorna E Clarson, Alice Faux-Nightingale, Tina Hadley-Barrows, John Haines, Rosie Harrison, Toby Helliwell, Samantha L Hider, Clare Jinks, Natalie Knight, Christian D Mallen, Victoria K Welsh, Claire Burton

**Affiliations:** Primary Care Centre Versus Arthritis, School of Medicine, Keele University, ST5 5BG, UK; Haywood Academic Rheumatology Centre, Midlands Partnership University NHS Foundation Trust, Haywood Hospital, Staffordshire, ST6 7AG, UK

**Keywords:** primary care, elective care, advice and guidance, health inequalities

## Abstract

**Objective:** to examine trends and variation in the use of Advice and Guidance (A&G) compared with direct referrals in primary care, and to assess potential disparities across population groups.

**Design:** Observational study using routinely collected electronic health records

**Setting:** Clinical Practice Research Datalink (CPRD) Aurum, 2015-2023.

**Main Outcome Measures:** annual prevalence of A&G and direct referrals, stratified by age, gender, deprivation, locality, and ethnicity. Mapping of clinical codes was used to determine target specialities for A&G. The proportion of individuals recorded with A&G and a direct referral within ±4 months was calculated.

**Results:** Between 2015-2023, 671,894 patients (4%; 59% female) had A&G recorded and 9.7 million (59%; 46% female) had a direct referral. A&G use increased 19-fold from (0.10% to 1.97%), doubling between 2019 and 2020 during the COVID-19 pandemic. Direct referral rates fell from 23%-25% pre-pandemic to 18% in 2020 before recovering to 24% by 2023. Cardiology (21%), Dermatology (7%) and Ear, Nose and Throat (5%) were the most common specialties linked to A&G. Most patients receiving A&G (86%) also had a direct referral within ±4 months. Inequities were evident: A&G use was higher among older, white, and less deprived patients, while minority ethnic and more deprived groups had slower recovery of direct referral rates post-pandemic.

**Conclusion:** A&G use has increased substantially since 2015, accelerated by the pandemic and maintained after, but has not displaced direct referrals. Instead, direct referral often precedes A&G, raising questions about efficiency and equity. The system appears to benefit older, white, and less deprived individuals while minority ethnic and more deprived groups remain disadvantaged. Policy should prioritise addressing these disparities and evaluate whether A&G reduces unnecessary referrals or delays access to specialist care.

**What is already known on this topic:**

– Advice & Guidance (A&G) was introduced to support outpatient reform and manage NHS waiting lists.
– Evidence on its usage patterns and equity across patient groups is limited.

**What this study adds:**

– A&G use increased 19-fold between 2015 and 2023, particularly post-pandemic.
– Disparities were identified with higher uptake in older, white and less deprived groups.
– Direct referrals have recovered to pre-pandemic levels, suggesting A&G is not replacing traditional pathways.

## Introduction

Referral is an important part of a primary care clinician’s role and describes a process that has a direct consequence on the patient experience of care as well as costs to the healthcare system. Referrals from primary care to secondary or specialist care may be made to establish a diagnosis, initiate specialist management, access an investigation not available in primary care, or seek a second opinion. Referral usually involves a transfer of clinical responsibility and can be a complex area of decision making, balancing the GP role as both patient advocate and NHS gatekeeper.[1–3]

In the UK National Health Service (NHS), referrals for elective (non-emergency) specialist care are usually made through the Choose & Book service (via the electronic Referral System [e-RS]), where the options include ‘Advice & Guidance’ (A&G) or ‘Refer/Advice.’ Refer/Advice allows the user to make a ‘direct referral’ to a particular specialty.[4] Established in 2015, A&G is a two-way dialogue delivered synchronously (voice call) or asynchronously (electronic communication) that enables primary care to seek specialist input into a patient’s care through formal recorded means. These two elective referral options are the focus of this study.

The use of A&G has been identified as a crucial part of NHS outpatient transformation to address the elective care backlog [5,6] with potential to shorten patient journeys through the system,[7,8] enable prompt appropriate management of conditions requiring specialist advice,[9] and reduce the burden on primary care that is exacerbated by long waits for specialist opinions.[10] The literature suggests that A&G can be efficient and effective in particular settings, but key aspects for service development include an understanding of suitable cases, equitable access, collaborative working across the health system, consideration of resource implications, and that existing A&G services may still yield high outpatient appointments with relatively few requests ending with expedited patient journeys (e.g. investigation requests and direct advice).[4,11–13]

However, it is unclear how and for which patient groups A&G is currently used and whether there is variation in its use by sociodemographic characteristics. Whilst the use of A&G has been identified as a crucial part of managing NHS waiting lists, there is little evidence to understand its effectiveness in reducing compound pressures whilst ensuring equity of use.

This first national study seeks to determine, within UK primary care, (i) the use of A&G over time in comparison to direct referrals; (ii) variation in A&G by socio-demographic characteristics; (iii) rates of A&G by specialty; and (iv) the percentage of A&G which end up with a direct referral.

## Materials and Methods

### Study Design and Setting

Electronic health record data were sourced from Clinical Practice Research Datalink (CPRD) Aurum, an anonymised UK primary care database covering 16.2 million individuals (∼24% UK population) from 1,784 current and historic general practices, primarily in England.[14]

The study was approved by the CPRD Research Data Governance (ref 24_004022) and used the June 2024 data release.[9] The approved protocol was made available to reviewers of this manuscript.

### Study Population

All individuals, regardless of age, with A&G or direct referral coded in their primary care record between 2015-2023 comprised the numerator population. Code lists for A&G and direct referrals are available here: https://doi.org/10.21252/t55f-vr93

Fast track referral for suspected cancer, and emergency referrals, or referral with an intention to admit, were excluded. The denominator population was the total registered population at the start of each calendar year.

### Covariates

Covariates included age at date of A&G request/referral, gender (as recorded in CPRD), geographical region, ethnicity based on the CPRD derived ethnicity algorithm,[15] neighbourhood deprivation (Index of Multiple Deprivation 2019 linked to patient postcode) categorised at the quintile scores, and Clinical Commissioning Group (CCG) pseudonym. CCGs were clinically-led organisations within the NHS that superseded primary care trusts in planning, assessing and purchasing healthcare services for 106 localities across England,[16] and were superseded by Integrated Care Boards (ICBs) on 1 July 2022. Given the use of both CCG and ICB localities during the study timeframe, we have used the term “*commissioning localities*”.

### Mapping A&G Records with Symptom/Diagnosis Codes

A mapping exercise allowed target specialities to be determined using coded events in the 14 days prior to and on the date of an A&G record. Using a random sample of 500,000 individuals with direct referral recorded in 2023 (categorised to e-RS referral specialties; https://doi.org/10.21252/t55f-vr93),[17] the most common symptom/diagnosis codes recorded 0-14 days before a direct referral were matched to symptom/diagnosis codes recorded 0-14 days before A&G where specialty was not recorded. Likely specialties were assigned to A&G records using the closest recorded code with the highest frequency specialty recorded for that code. Following review of the most frequent direct referral specialties by GP co-authors (CB, TH & VKW) referrals to physiotherapy, private practitioners, health promotion, occupational health, speech/language therapy, or orthotics/prosthetics were not included as likely specialties/services for A&G.

### Comparing prevalence of direct referrals to NHS e-RS Open Data Dashboard

In order to validate referral rates obtained from CPRD Aurum, referral data recorded between 30/12/2019 - 08/09/2024 were obtained from the NHS e-Referral Service (e-RS) Open Data Dashboard.[17] Referrals were included in annual totals if categorised as “*routine*” or “*urgent*” and excluded if categorised as 2 week wait and/or cancer referrals. Annual mid-year population estimates for England between 2020-2023 were obtained from the Office for National Statistics (ONS) and used as the denominator population to calculate rates for the e-RS referrals data.[18]

### Statistical Analysis

Annual prevalence rates for A&G, direct referrals and NHS e-RS Open Data Dashboard were calculated as a percentage of the registered population having at least 1 A&G (or referral, as appropriate) recorded during the year.

Trends of A&G and direct referrals were analysed overall and stratified by age, gender, deprivation, ethnicity, geographical region and commissioning localities to assess variations over time.

The percentage of individuals recorded with A&G also recorded with a direct referral in the 4 months (122 days; agreed by clinical consensus) i) before (−122 to 0 days) and ii) after (1 to 122 days) A&G record were derived and compared by the covariates (excluding those with less than four months of follow-up), descriptively and by using multinomial logistic regression (relative risk ratio [RRR] and adjusted RRR [aRRR] presented with 95% confidence intervals [CI]). Age, gender, region, ethnicity, deprivation and commissioning locality were included as covariates in adjusted models. A sensitivity analysis expanding to 12 months before and after A&G record was also performed (excluding those with less than 12 months of follow-up).

Absence of a code for A&G or direct referral was assumed to indicate no consultation having occurred. All included individuals had year of birth (to calculate age), consultation year and gender recorded.

Individuals without data for ethnicity were recorded as “*unknown*” and as “*missing*” for deprivation. Data management and statistical analyses were performed using Stata MP version 19.5 (StataCorp, USA).

This study meets all five of the CODE-EHR minimum framework standards for the use of structured healthcare data in clinical research, with one preferred criteria also met (Ethics and Governance). Full details are provided in the completed CODE-EHR checklist (Supplementary Table S1).[19]

### Patient and Public Involvement

Patients and the public were involved at all stages of this study. A public contributor (JH) was a co-applicant on the grant and attended monthly study meetings. Our dedicated Patient and Public Involvement (PPI) group advised on study design, ensuring that our focus on disparities in A&G use reflected patient priorities. They highlighted issues such as digital literacy and access to transport, which informed our interpretation of inequities. PPI members reviewed the findings and contributed to the framing of recommendations around efficiency and equity. They meet every 3 months to ensure they remain connected with the study and able to provide insight and input on a regular basis and will support dissemination through plain-language summaries. We recognise that our PPI group was not fully representative of all ethnic and socioeconomic backgrounds affected by disparities, and future work will prioritise broader inclusion.

## Results

We analysed data on 16.3 million registered patients. Between 2015-2023, 4% had A&G recorded and 59% had a direct referral (Table 1). A&G was more commonly recorded in older patients and females with direct referrals recorded more commonly in males, and both elective care pathways more commonly recorded for those of white ethnicity and least deprived individuals (Table 1).

**Table 1.**
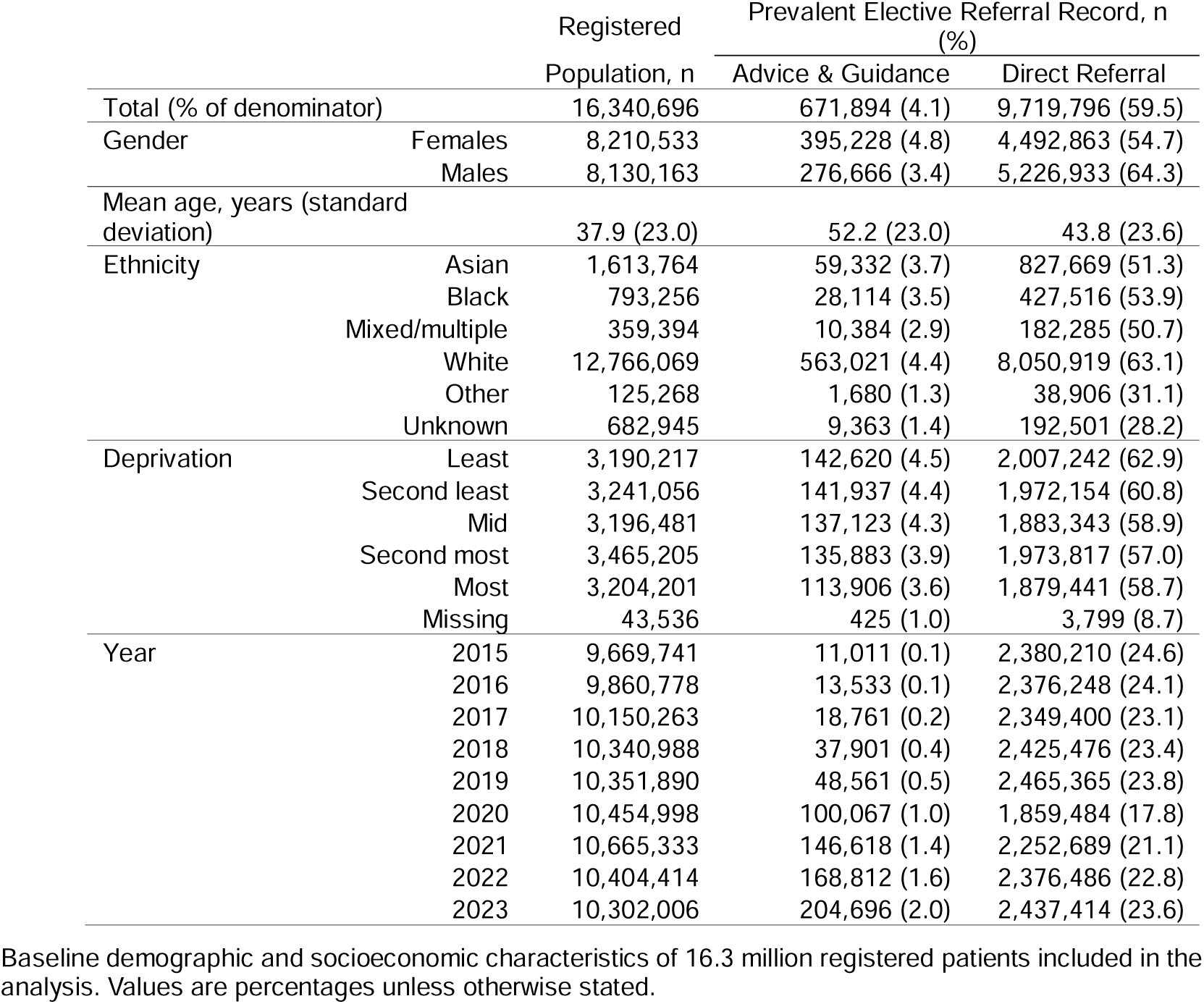
Characteristics of the study population (2015 to 2023)

|  |  | Registered | Prevalent Elective Referral Record, n (%) |  |
| --- | --- | --- | --- | --- |
|  |  | Population, n | Advice & Guidance | Direct Referral |
| Total (% of denominator) |  | 16,340,696 | 671,894 (4.1) | 9,719,796 (59.5) |
| Gender | Females | 8,210,533 | 395,228 (4.8) | 4,492,863 (54.7) |
|  | Males | 8,130,163 | 276,666 (3.4) | 5,226,933 (64.3) |
| Mean age, years (standard deviation) |  | 37.9 (23.0) | 52.2 (23.0) | 43.8 (23.6) |
| Ethnicity | Asian | 1,613,764 | 59,332 (3.7) | 827,669 (51.3) |
|  | Black | 793,256 | 28,114 (3.5) | 427,516 (53.9) |
|  | Mixed/multiple | 359,394 | 10,384 (2.9) | 182,285 (50.7) |
|  | White | 12,766,069 | 563,021 (4.4) | 8,050,919 (63.1) |
|  | Other | 125,268 | 1,680 (1.3) | 38,906 (31.1) |
|  | Unknown | 682,945 | 9,363 (1.4) | 192,501 (28.2) |
| Deprivation | Least | 3,190,217 | 142,620 (4.5) | 2,007,242 (62.9) |
|  | Second least | 3,241,056 | 141,937 (4.4) | 1,972,154 (60.8) |
|  | Mid | 3,196,481 | 137,123 (4.3) | 1,883,343 (58.9) |
|  | Second most | 3,465,205 | 135,883 (3.9) | 1,973,817 (57.0) |
|  | Most | 3,204,201 | 113,906 (3.6) | 1,879,441 (58.7) |
|  | Missing | 43,536 | 425 (1.0) | 3,799 (8.7) |
| Year | 2015 | 9,669,741 | 11,011 (0.1) | 2,380,210 (24.6) |
|  | 2016 | 9,860,778 | 13,533 (0.1) | 2,376,248 (24.1) |
|  | 2017 | 10,150,263 | 18,761 (0.2) | 2,349,400 (23.1) |
|  | 2018 | 10,340,988 | 37,901 (0.4) | 2,425,476 (23.4) |
|  | 2019 | 10,351,890 | 48,561 (0.5) | 2,465,365 (23.8) |
|  | 2020 | 10,454,998 | 100,067 (1.0) | 1,859,484 (17.8) |
|  | 2021 | 10,665,333 | 146,618 (1.4) | 2,252,689 (21.1) |
|  | 2022 | 10,404,414 | 168,812 (1.6) | 2,376,486 (22.8) |
|  | 2023 | 10,302,006 | 204,696 (2.0) | 2,437,414 (23.6) |

Baseline demographic and socioeconomic characteristics of 16.3 million registered patients included in the analysis. Values are percentages unless otherwise stated.

### Annual Prevalence of A&G and Direct Referrals

A&G use rose almost 20-fold from 0.1% in 2015 to 2.0% in 2023, with the sharpest rise during the COVID-19 pandemic (Figure 1). Direct referrals fell from ∼24% pre-pandemic to 18% in 2020, before recovering to 24% by 2023 (Figure 1). The ratio of direct referral prevalence to A&G prevalence fell from 235:1 in 2015 to 12:1 in 2023.

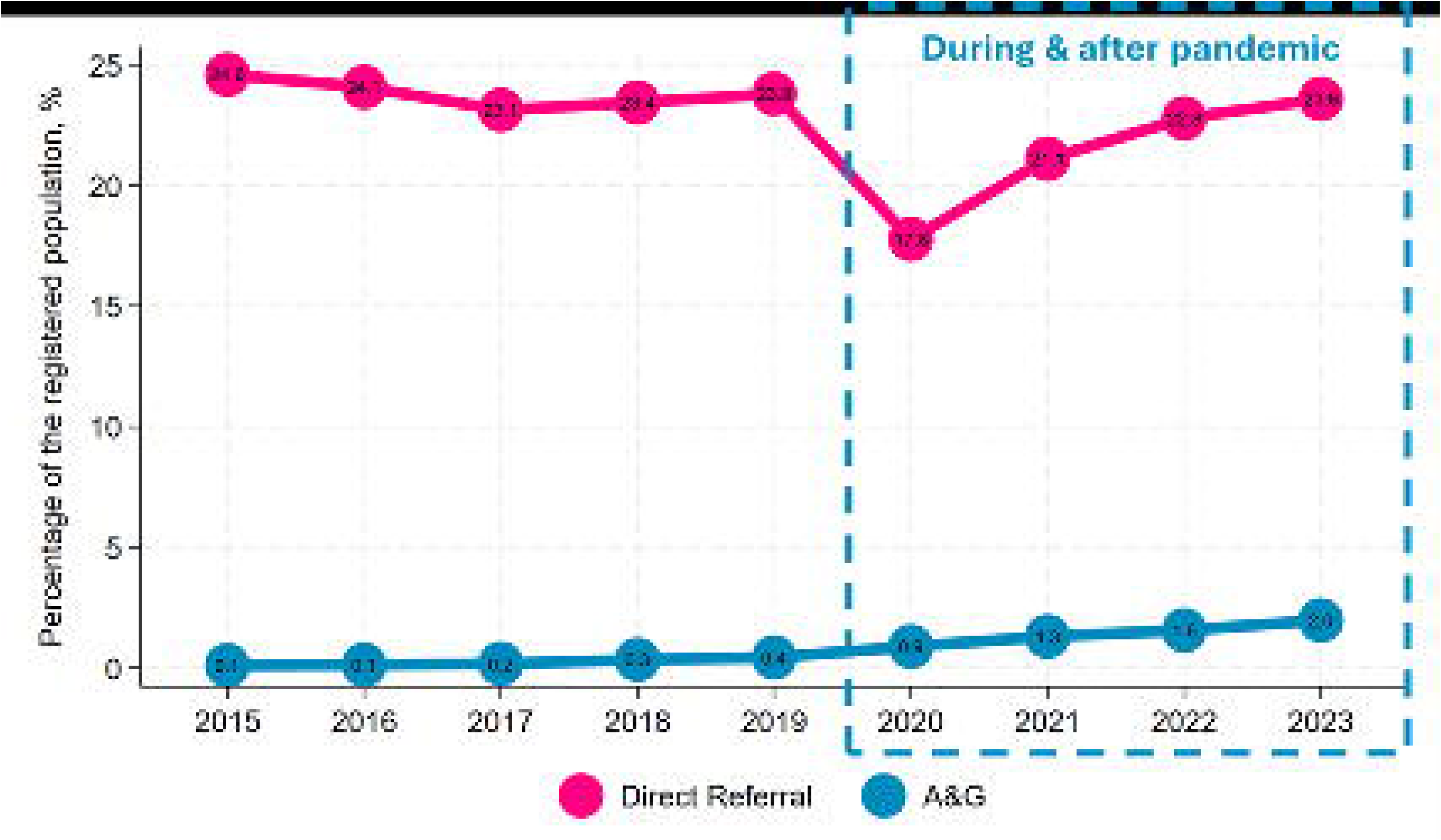

### Rates of A&G by Age, Gender, Ethnicity and Deprivation

Figure 2 depicts trends in prevalence of A&G and direct referrals over time stratified by age, gender, ethnicity, and deprivation level. Although A&G is more prevalent in males over time, a higher percentage of females were recorded with A&G due to males experiencing more episodes. A&G prevalence increased with age for both genders from age 25, peaking in the elderly (75–84 years; Figure 2). A sharp rise is observed from 2020 to 2023 (green dashed lines), particularly in older adults (Supplementary Table S2).

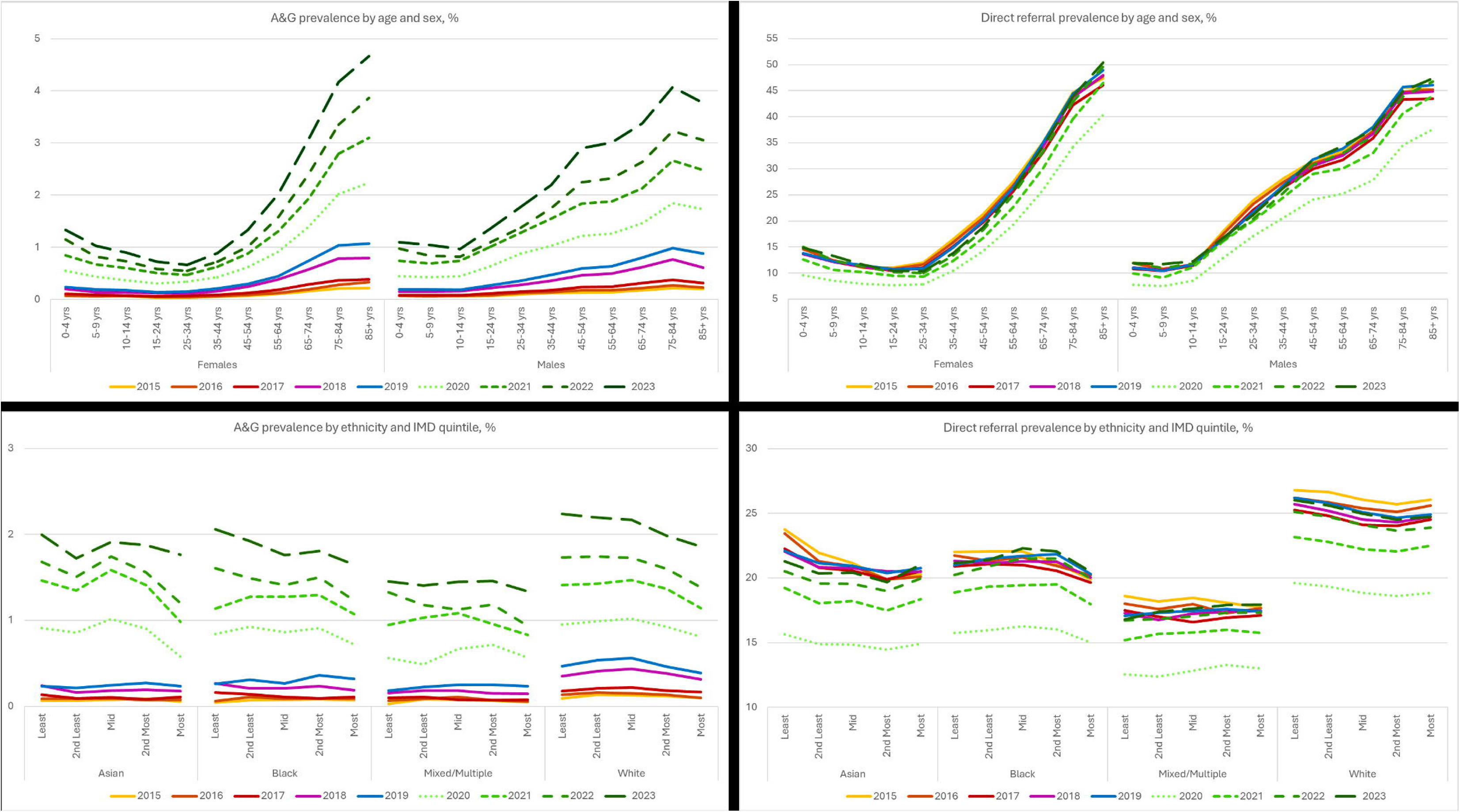

White individuals had higher A&G prevalence than Asian, Black, and Mixed/Multiple ethnic groups (Figure 2). Rates of A&G doubled between 2019-2020 for all ethnic groups, likely due to the COVID-19 pandemic. Disparity in use was observed with higher A&G prevalence in less deprived quintiles across all ethnic groups from 2020 onwards (Supplementary Table S2).

### Rates of Direct Referral by Age, Gender, Ethnicity and Deprivation

Direct referrals increased with age from 25 years in females and 15 years in males, peaking in those aged 75–84 years, with slightly higher referral prevalence in males than females for most age groups (Figure 2). Prevalence rates of direct referrals dropped in 2020 (lightest green dotted line; Figure 2), with referral rates increasing in each following year to pre-pandemic levels by 2023 (Supplementary Table S3).

Direct referrals decreased with increasing deprivation across ethnic groups in the years preceding the COVID-19 pandemic (2015-2019) and post pandemic (2022-2023; Figure 2). White patients had the highest referral prevalence, with the mixed/multiple ethnic group having the lowest rates. In 2020-2021 (lighter green dashed lines; Figure 2), referral rates dipped significantly, particularly among more deprived and ethnic minority groups (Supplementary Table S3).

### Recent Variation in Rates of A&G and Direct Referrals by Covariates

Focusing on 2023 (due to the highest prevalence of A&G and recovery of direct referral rates to pre-pandemic levels) a greater prevalence of A&G and direct referrals were observed for males, older adults, white ethnicity, and less deprived individuals (Table 2). Wide variation was observed for commissioning locality use of A&G within each region of England, particularly in the North (Figure 3; Supplementary Table S4). However, higher A&G activity was not associated with lower direct referral rates (correlation coefficient = 0.01; P=0.925), indicating that A&G use does not necessarily translate into reduced direct referral (Supplementary Table S4).

**Table 2.**
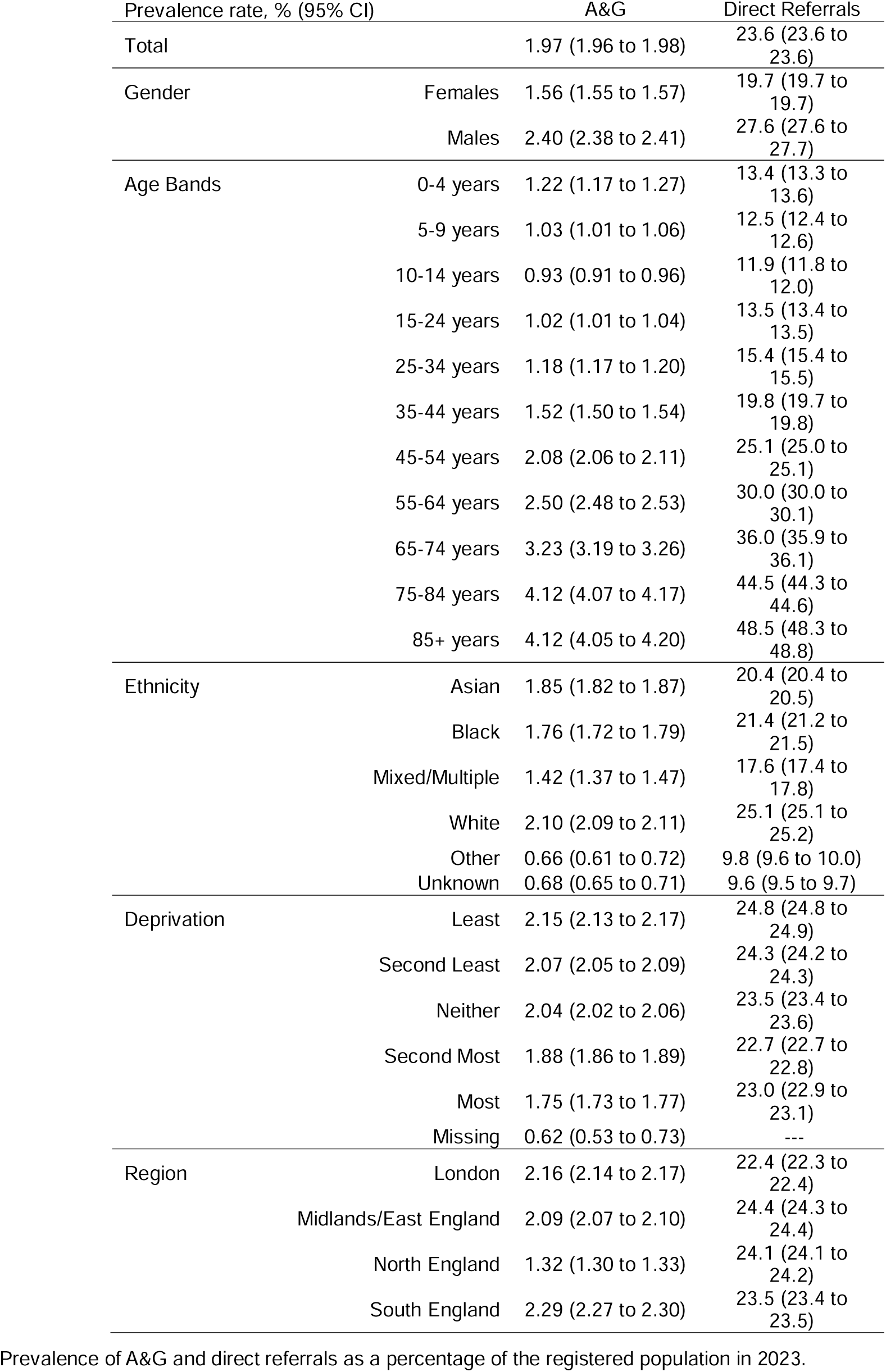
Annual prevalence of A&G and direct referrals (2023)

| Prevalence rate, % (95% CI) |  | A&G | Direct Referrals |
| --- | --- | --- | --- |
| Total |  | 1.97 (1.96 to 1.98) | 23.6 (23.6 to 23.6) |
| Gender | Females | 1.56 (1.55 to 1.57) | 19.7 (19.7 to 19.7) |
|  | Males | 2.40 (2.38 to 2.41) | 27.6 (27.6 to 27.7) |
| Age Bands | 0-4 years | 1.22 (1.17 to 1.27) | 13.4 (13.3 to 13.6) |
|  | 5-9 years | 1.03 (1.01 to 1.06) | 12.5 (12.4 to 12.6) |
|  | 10-14 years | 0.93 (0.91 to 0.96) | 11.9 (11.8 to 12.0) |
|  | 15-24 years | 1.02 (1.01 to 1.04) | 13.5 (13.4 to 13.5) |
|  | 25-34 years | 1.18 (1.17 to 1.20) | 15.4 (15.4 to 15.5) |
|  | 35-44 years | 1.52 (1.50 to 1.54) | 19.8 (19.7 to 19.8) |
|  | 45-54 years | 2.08 (2.06 to 2.11) | 25.1 (25.0 to 25.1) |
|  | 55-64 years | 2.50 (2.48 to 2.53) | 30.0 (30.0 to 30.1) |
|  | 65-74 years | 3.23 (3.19 to 3.26) | 36.0 (35.9 to 36.1) |
|  | 75-84 years | 4.12 (4.07 to 4.17) | 44.5 (44.3 to 44.6) |
|  | 85+ years | 4.12 (4.05 to 4.20) | 48.5 (48.3 to 48.8) |
| Ethnicity | Asian | 1.85 (1.82 to 1.87) | 20.4 (20.4 to 20.5) |
|  | Black | 1.76 (1.72 to 1.79) | 21.4 (21.2 to 21.5) |
|  | Mixed/Multiple | 1.42 (1.37 to 1.47) | 17.6 (17.4 to 17.8) |
|  | White | 2.10 (2.09 to 2.11) | 25.1 (25.1 to 25.2) |
|  | Other | 0.66 (0.61 to 0.72) | 9.8 (9.6 to 10.0) |
|  | Unknown | 0.68 (0.65 to 0.71) | 9.6 (9.5 to 9.7) |
| Deprivation | Least | 2.15 (2.13 to 2.17) | 24.8 (24.8 to 24.9) |
|  | Second Least | 2.07 (2.05 to 2.09) | 24.3 (24.2 to 24.3) |
|  | Neither | 2.04 (2.02 to 2.06) | 23.5 (23.4 to 23.6) |
|  | Second Most | 1.88 (1.86 to 1.89) | 22.7 (22.7 to 22.8) |
|  | Most | 1.75 (1.73 to 1.77) | 23.0 (22.9 to 23.1) |
|  | Missing | 0.62 (0.53 to 0.73) | --- |
| Region | London | 2.16 (2.14 to 2.17) | 22.4 (22.3 to 22.4) |
|  | Midlands/East England | 2.09 (2.07 to 2.10) | 24.4 (24.3 to 24.4) |
|  | North England | 1.32 (1.30 to 1.33) | 24.1 (24.1 to 24.2) |
|  | South England | 2.29 (2.27 to 2.30) | 23.5 (23.4 to 23.5) |

Prevalence of A&G and direct referrals as a percentage of the registered population in 2023.

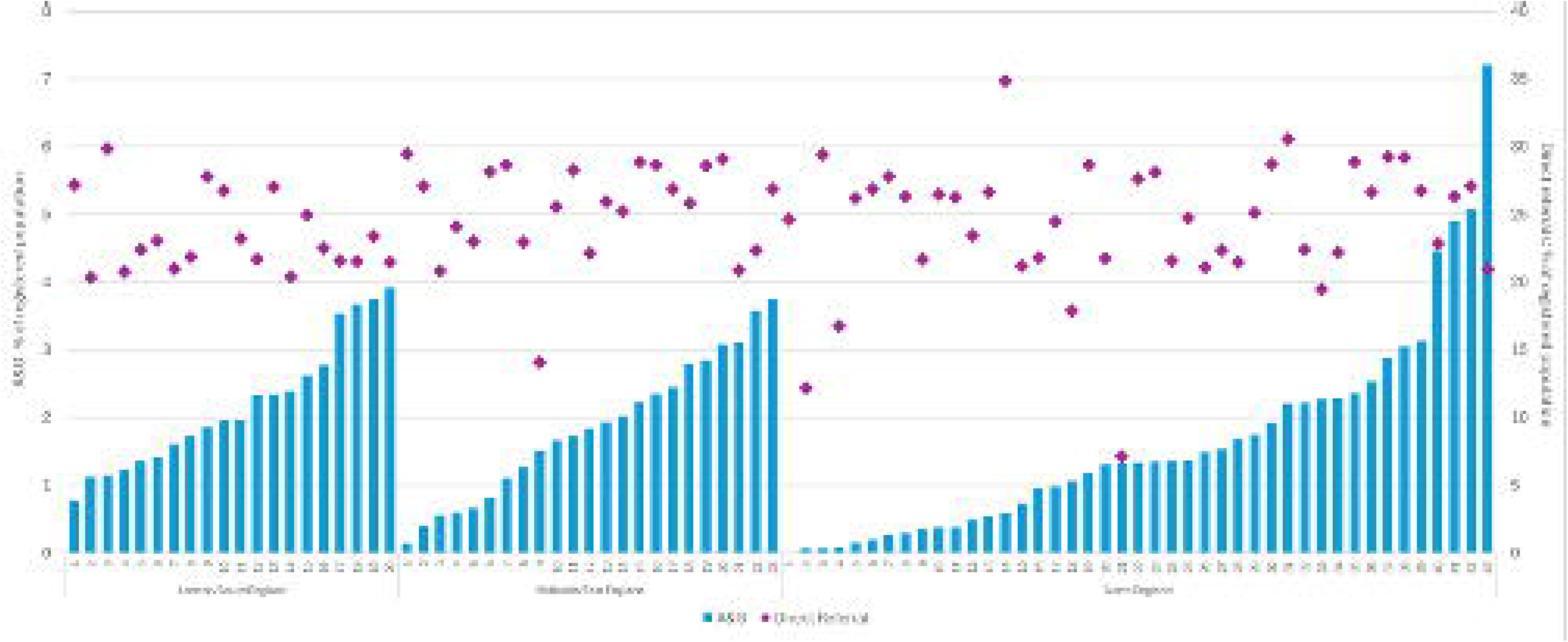

### Proportion of Individuals with A&G and Direct Referrals Recorded

Among patients recorded with A&G, 86% also had a direct referral within ±4 months (Table 3). Direct referral more often preceded A&G (69%) than followed it (17%), indicating that A&G is frequently used after referral rather than as a substitute or precursor (Table 3). This challenges the assumption that A&G typically precedes referral. Disparities were evident: A&G was more commonly used by older, white, and less deprived patients, while minority ethnic groups were more likely to have direct referral after A&G, suggesting potential delays in access to specialist care for these populations (Table 3). These patterns persisted in sensitivity analyses using a ±12-month window (Supplementary Table S5).

**Table 3.**
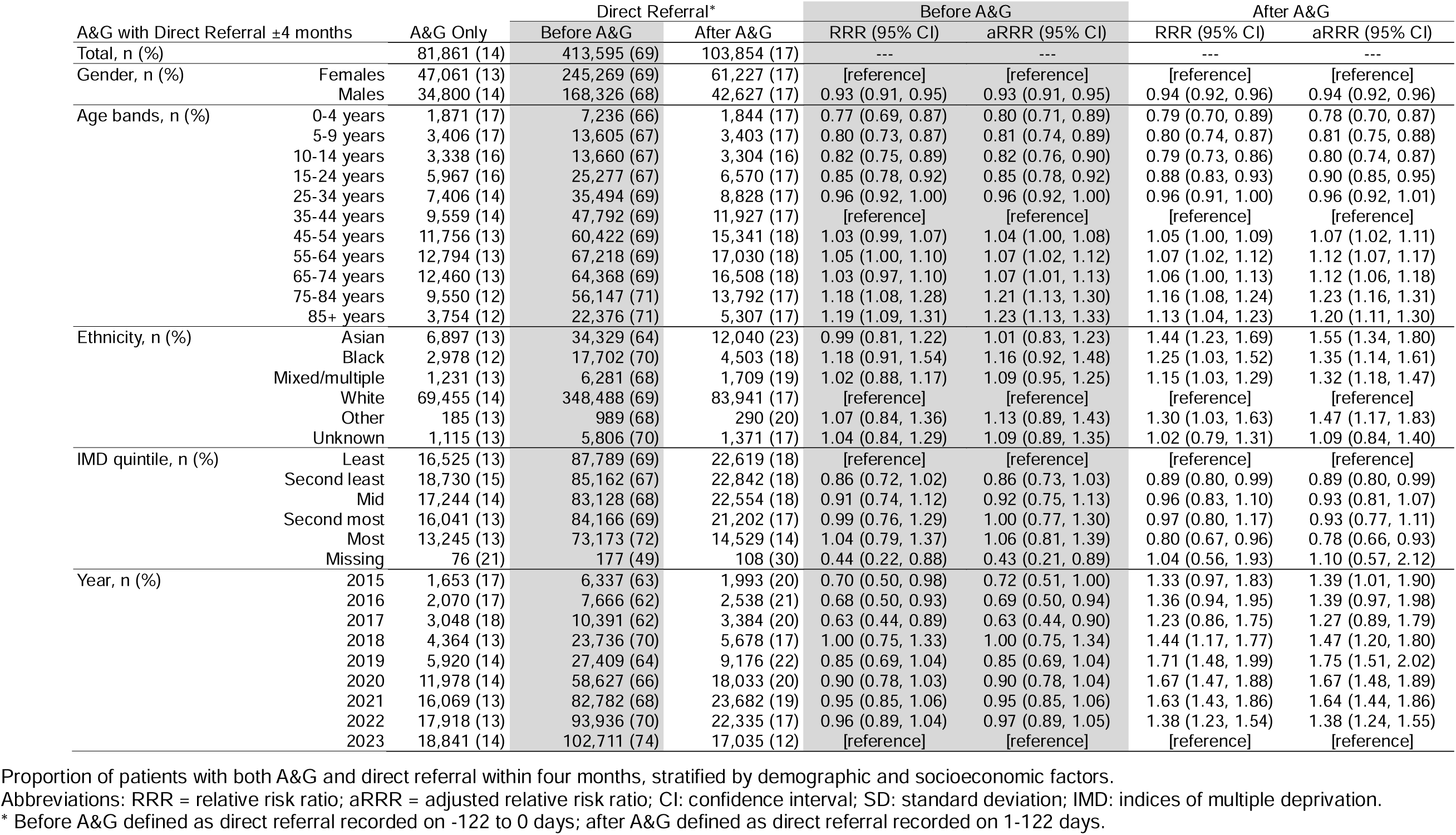
Association between A&G and Direct Referral within ±4 months.

### Mapping

Of 162,787 individuals recorded with their first A&G record in 2023, 2% had a specialty recorded in 0-14 days prior to A&G. Matching consultation codes recorded for direct referrals resulted in an additional 59% (61% total) attributed to a specialty with Cardiology, Dermatology, and Ear, Nose and Throat the most frequent (Figure 4).

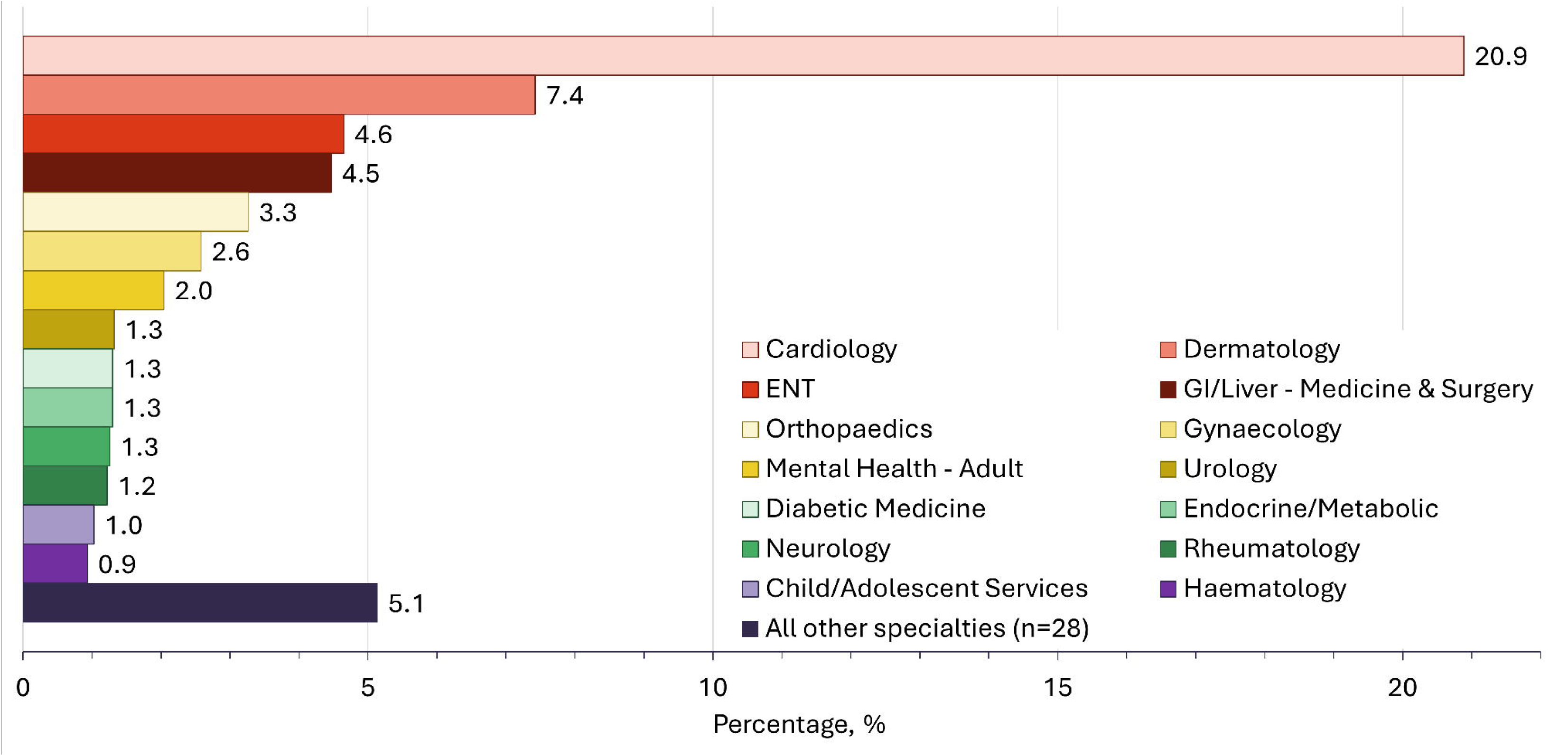

### Comparison to e-RS Dashboard Data

Rates of direct referral in CPRD Aurum were comparable to rates calculated using data from the NHS e-RS Open Data Dashboard (Supplementary Table S6).

## Discussion

We observed an almost twentyfold increase in the use of A&G during and following the COVID-19 pandemic, representing a systematic shift in how specialist input is accessed.[6] However, this expansion did not displace direct referrals, which returned to pre-pandemic levels by 2023. Most patients receiving A&G were also referred within four months, suggesting the service often adds a step rather than substitutes for referral.

Inequities were clear: A&G was more often used for older, white, and less deprived patients, while minority ethnic and more deprived groups showed slower recovery of direct referral access after the pandemic. At locality level, patterns varied widely, and higher A&G use was not associated with lower referral rates raising questions about the efficiency of the pathway and whether it genuinely empowers GPs to manage more care in the community.[5]

### Comparison to Literature

Our findings align with national policy initiatives during the COVID-19 pandemic,[6] which promoted A&G to maintain specialist input while reducing face-to-face outpatient activity and attempt to reduce pressures on the elective healthcare system in the pandemic recovery. NHS England’s elective care recovery plan aimed to deliver 30% more elective activity by 2024/25 compared with pre-pandemic levels (2019/20), partly through wider A&G use. [20] While we could not calculate A&G-to-outpatient ratios, our results suggest substantial progress towards this target.

However, the persistence of high onward referral rates indicates that A&G is not yet fulfilling its intended role as a substitute for direct referral; instead, it appears to add an additional step of unknown benefit in many patient journeys.

The socioeconomic and ethnic disparities we observed mirror wider evidence that the COVID-19 pandemic exacerbated existing health inequalities in England, particularly for deprived and minority ethnic groups.[21] The King’s Fund subsequently conducted research into how these disparities were being addressed, highlighting barriers such as transport, employment constraints, and lower health literacy.[22] Our research suggests inequities may also arise earlier, at the point of referral itself, with A&G disproportionately applied to certain groups before a referral to secondary care is made.

NHS England instructed trusts in 2020 to disaggregate waiting lists by ethnicity and deprivation, but reducing inequities will require action upstream, including the point of referral. Our findings support calls for more inclusive approaches to outpatient transformation,[7,20] ensuring that initiatives such as A&G do not unintentionally widen gaps in access.

### Strengths and Limitations

This is the first national, population-based study to explore trends and variations in A&G versus direct referrals from primary care in the NHS. The large dataset and longitudinal design provide a unique opportunity to examine the impact of system-wide changes, including those accelerated by the pandemic.

Limitations include the potential for misclassification of A&G and referral coding, though any such errors are likely to be non-differential. Uptake of alternative tele-medicine providers may not be consistently coded in electronic health records meaning some specialist input is unobserved.

Automation of A&G recording in existing electronic health record systems could enhance clinician usability and record-keeping. Our analysis is also descriptive and cannot establish causality between service design, sociodemographic characteristics, and referral outcomes. Lastly, recent policy changes incentivising A&G coding were too recent to be captured in our data.[23]

### Policy Implications

For A&G to support NHS priorities—reducing elective backlogs and moving care into the community— three actions are required:

– *<u>Efficiency</u>*: evaluate whether A&G prevents unnecessary referrals or simply delays them, including assessment of patient outcomes and system costs;
– *<u>Equity</u>*: routinely monitor A&G by deprivation and ethnicity, and support practices serving disadvantaged populations to ensure that referral pathways do not reinforce inequalities;
– *<u>Primary care capacity</u>*: invest in GP training, coding support, and digital infrastructure so A&G enables community management rather than becoming an additional administrative step.

Without these measures, A&G risks functioning as a bottleneck in the referral process rather than a tool for transformation aligned to delivery of the NHS ten-year plan.[5]

## Conclusion

A&G use has risen sharply across the NHS, but this has not reduced direct referrals. Instead, A&G appears to be layered onto existing pathways, with potential inefficiencies and risks of widening inequities. Older, white, and less deprived populations are disproportionately accessing A&G, while minority ethnic groups are more often subject to direct referral following A&G.

If A&G is to contribute meaningfully to the NHS Long-Term Plan to shift care into the community and reduce elective care backlogs,[24] it must be redesigned and resourced to improve efficiency and equity, ensuring it contributes to shorter, fairer, and more effective patient pathways.

## Copyright/License for Publication

The Corresponding Author has the right to grant on behalf of all authors and does grant on behalf of all authors, a worldwide licence to the Publishers and its licensees in perpetuity, in all forms, formats and media (whether known now or created in the future), to

1. publish, reproduce, distribute, display and store the Contribution,
2. translate the Contribution into other languages, create adaptations, reprints, include within collections and create summaries, extracts and/or, abstracts of the Contribution,
3. create any other derivative work(s) based on the Contribution,
4. to exploit all subsidiary rights in the Contribution,
5. the inclusion of electronic links from the Contribution to third party material where-ever it may be located; and,
6. licence any third party to do any or all of the above.

## Availability of Data and Materials

Data may be obtained from a third party and are not publicly available. The data were obtained from the Clinical Practice Research Datalink. Clinical Practice Research Datalink data governance does not allow us to distribute patient data to other parties. Researchers may apply for data access at http://www.CPRD.com/.

Code lists are openly available at https://doi.org/10.21252/t55f-vr93

## Ethics Approval

CPRD has Health Research Authority approval to support research using anonymised patient data. This study was approved by the CPRD Research Data Governance Process (ref 24_0040223; protocol made available to this manuscript’s reviewers). Under CPRD’s ethical approval from the UK Health Research Authority to support research using anonymised patient data, individual patient consent is not required as patients contributing data to CPRD cannot be identified from the data made available to researchers.

Data from CPRD adheres to the ethical principles outlined in the Declaration of Helsinki. Code lists are available via https://doi.org/10.21252/t55f-vr93

## Transparency

The lead author (KJM) affirms that the manuscript is an honest, accurate, and transparent account of the study being reported; that no important aspects of the study have been omitted; and that any discrepancies from the study as originally planned (and, if relevant, registered) have been explained.

## Authors’ Contributions

All authors were involved in the writing - original draft and writing – review & editing of the manuscript. Attributed author roles are summarised below. *Conceptualisation and funding acquisition:* KJM, KPJ, JB, RB, AFN, JH, RH, SLH, TH, CJ, CDM, VKW, CB *Data curation, formal analysis and validation:* KJM, JB *Methodology and project administration:* KJM, JB *Supervision*: KPJ, VKW, CB *Visualisation*: KJM

## Data Availability

Data may be obtained from a third party and are not publicly available. The data were obtained from the Clinical Practice Research Datalink. Clinical Practice Research Datalink data governance does not allow us to distribute patient data to other parties. Researchers may apply for data access at http://www.CPRD.com/.
Code lists are openly available at https://doi.org/10.21252/t55f-vr93

## Acknowledgments

CPRD: This study is based in part on data from the Clinical Practice Research Datalink obtained under licence from the UK Medicines and Healthcare products Regulatory Agency. The data is provided by patients and collected by the NHS as part of their care and support. The interpretation and conclusions contained in this study are those of the authors alone. PPI/Advisory group: The authors would especially like to thank the members of the Research User Group members involved BADGER, particularly John Haines as public co-applicant on the grant, for their valuable contributions to this study. We are also thankful to Keele PPI team for their support of the public contributors. The authors would also like to thank the NIHR ARC West Midlands and the Study Oversight Committee for their support.

## References

1. The King’s Fund. The quality of GP diagnosis and referral. Diagnosis and referral. kingsfund.org.uk. 2010.

2. Bi YN, Liu YA. GPs in UK: From Health Gatekeepers in Primary Care to Health Agents in Primary Health Care. Risk Manag Healthc Policy [Internet]. 2023 [cited 2025 Jul 17];16:1929. Available from: https://pmc.ncbi.nlm.nih.gov/articles/PMC10518152/

3. Royal College of General Practitioners. Quality patient referrals: Right service, right time. Quality patient referrals - Right Service, right time - February 2018 [Internet]. rcgp.org.uk. 2018 [cited 2025 Jul 17]. Available from: https://www.rcgp.org.uk/representing-you/policy-areas/referral-management

4. NHS Digital. Advice and guidance toolkit for the NHS e-Referral Service (e-RS) - NHS England Digital [Internet]. 2022 [cited 2025 Mar 17]. Available from: https://digital.nhs.uk/services/e-referral-service/document-library/advice-and-guidance-toolkit

5. Fit for the future: 10 Year Health Plan for England (accessible version) - GOV.UK [Internet]. [cited 2025 Jul 15]. Available from: https://www.gov.uk/government/publications/10-year-health-plan-for-england-fit-for-the-future/fit-for-the-future-10-year-health-plan-for-england-accessible-version

6. NHS England. Elective Care Transformation Programme [Internet]. 2023 [cited 2025 Mar 17]. Available from: https://www.england.nhs.uk/elective-care/

7. NHS Englandl» Reforming elective care for patients [Internet]. [cited 2025 Jul 15]. Available from: https://www.england.nhs.uk/publication/reforming-elective-care-for-patients/

8. Anderson KN, Warren N, Duddy M, McKean P, Miller JAL. Delivering an advice and guidance service in neurology. Pract Neurol [Internet]. 2022 Jun 1 [cited 2025 Mar 17];22(3):209–12. Available from: https://pn.bmj.com/content/22/3/209

9. Moghaddas F, Tsiougkos N, Grammatikos A, Bright PD, Johnston S, Gompels M. COVID-19 vaccine allergy advice and guidance: The experience of a UK tertiary referral centre. World Allergy Organ J [Internet]. 2023 Jan 1 [cited 2025 Mar 17];16(1). Available from: https://pubmed.ncbi.nlm.nih.gov/36644019/

10. Radnaeva I, Muthusami A, Ward S. NHS advice and guidance – improving outpatient flow and patient care in general surgery. The Surgeon. 2023 Oct 1;21(5):e258–62.

11. Royal College of General Practitioners. Advice and guidance [Internet]. 2022 [cited 2025 Mar 17]. Available from: https://www.rcgp.org.uk/representing-you/policy-areas/advice-and-guidance

12. Pulse Today. Two thirds of GPs say ‘advice and guidance’ is blocking patients who really need a referral - Pulse Today [Internet]. 2023 [cited 2025 Mar 17]. Available from: https://www.pulsetoday.co.uk/news/workload/two-thirds-of-gps-say-advice-and-guidance-is-blocking-patients-who-really-need-a-referral/

13. The Patients Association. Understanding patient perspectives on improving GP referrals to secondary care through the use of specialist advice and guidance Patient panel workshops Final report on findings and recommendations [Internet]. 2022 [cited 2025 Mar 17]. Available from: https://pexlib.net/?237824

14. CPRD Aurum June 2024 dataset | CPRD [Internet]. [cited 2025 Mar 17]. Available from: https://www.cprd.com/doi/cprd-aurum-june-2024-dataset

15. Shiekh SI, Harley M, Ghosh RE, Ashworth M, Myles P, Booth HP, et al. Completeness, agreement, and representativeness of ethnicity recording in the United Kingdom’s Clinical Practice Research Datalink (CPRD) and linked Hospital Episode Statistics (HES). Popul Health Metr [Internet]. 2023 Dec 1 [cited 2025 Mar 12];21(1):1–13. Available from: https://pophealthmetrics.biomedcentral.com/articles/10.1186/s12963-023-00302-0

16. Hammond J, Coleman A, Checkland K. Enacting localist health policy in the English NHS: The ‘governing assemblage’ of clinical commissioning groups. J Health Serv Res Policy [Internet]. 2018 Jan 1 [cited 2025 Jul 15];23(1):49–56. Available from: https://pubmed.ncbi.nlm.nih.gov/29256272/

17. NHS e-Referral Service (e-RS) open data dashboard - NHS England Digital [Internet]. [cited 2025 Jun 3]. Available from: https://digital.nhs.uk/dashboards/ers-open-data

18. England population mid-year estimate - Office for National Statistics [Internet]. [cited 2025 Jun 3]. Available from: https://www.ons.gov.uk/peoplepopulationandcommunity/populationandmigration/populationestimates/timeseries/enpop/pop

19. Kotecha D, Asselbergs FW, Achenbach S, Anker SD, Atar D, Baigent C, et al. CODE-EHR best practice framework for the use of structured electronic healthcare records in clinical research. BMJ [Internet]. 2022 Aug 29 [cited 2025 Sep 4];378:51. Available from: https://www.bmj.com/content/378/bmj-2021-069048

20. National Audit Office. NHS England; NHS England’s management of elective care transformation programmes. 2025 [cited 2025 Jul 15]; Available from: https://www.nao.org.uk/reports/nhs-englands-management-of-elective-care-transformation-programmes/

21. Health inequalities in a nutshell | The King’s Fund [Internet]. [cited 2025 Jul 15]. Available from: https://www.kingsfund.org.uk/insight-and-analysis/data-and-charts/health-inequalities-nutshell

22. Tackling Health Inequalities On NHS Waiting Lists | The King’s Fund [Internet]. [cited 2025 Jul 15]. Available from: https://www.kingsfund.org.uk/insight-and-analysis/reports/health-inequalities-nhs-waiting-lists

23. NHS England_l_» Enhanced service specification – General Practice Requests for Advice and Guidance [Internet]. [cited 2025 Jul 15]. Available from: https://www.england.nhs.uk/publication/enhanced-service-specification-general-practice-requests-for-advice-and-guidance/

24. NHS England_l_» The NHS Performance Assessment Framework for 2025/26 [Internet]. [cited 2025 Jul 15]. Available from: https://www.england.nhs.uk/long-read/the-nhs-performance-assessment-framework-for-2025-26/

