## Supplementary Materials for "Trends and inequalities in Advice and Guidance versus direct referral in NHS primary care: a population-based study, 2015–2023"

Supplementary Tables

Table S1	CODE-EHR Framework: Best practice checklist to report on the use of structured electronic healthcare records in clinical research

Table S2	Prevalence of A&G by Sociodemographic Factors and Year, %

Table S3	Prevalence of Direct Referrals by Sociodemographic Factors and Year, %

Table S4	Mean Variation in A&G and Direct Referrals by Region: 2023

Table S5	Characteristics of those with/without Direct Referrals within ±12 months of A&G

Table S6	Comparison of Direct Referral Rates from CPRD Aurum to e-RS Dashboard

Table S1	CODE-EHR Framework: Best practice checklist to report on the use of structured electronic healthcare records in clinical research

Date of completion: 04/Sep/25
Study name: Trends and inequalities in Advice and Guidance versus direct referral in NHS primary care: a population-based study, 2015–2023

| **Item** | **Objective** | **Framework standards** | **Minimum information to provide** | **Lead Author acknowledgement** |
| --- | --- | --- | --- | --- |
| 1. Dataset construction and linkage | To provide an understanding of how the structured healthcare data were identified and used. | Minimum: Flow diagram of datasets used in the study, and description of the processes and directionality of any linkage performed, published within the research report or supplementary documents.  Preferred: Provided within a pre-published protocol or open-access document. | (a) State the source of any datasets used.  (b) Comment on how the observed and any missing data were identified and addressed, and the proportion observed for each variable. (c) Provide data on completeness of follow-up. (d) For linked datasets, specify how linkage was performed and the quality of linkage methods. | Select one option:  Minimum standard not met ☐ Minimum standard met ☒  Preferred standard met ☐ |
| 2. Data fit for purpose | To ensure transparency with the approach taken, with respect to coding of the structured healthcare data. | Minimum: Clear unambiguous statements on the process of coding in the methods section of the research report.  Preferred: Provided within a pre-published protocol or open-access document. | (a) Confirm origin, clinical processes, and the purpose of data. (b) Specify coding systems, clinical terminologies or classification used and their versions, and any manipulation of the coded data.  (c) Provide detail on quality assessment for data capture. (d) Outline potential sources of bias. | Select one option:  Minimum standard not met ☐ Minimum standard met ☒  Preferred standard met ☐ |
| 3. Disease and outcome definitions | To fully detail how conditions AND outcome events were defined, allowing other researchers to identify errors and repeat the process in other datasets. | Minimum: State what codes were used to define diseases, treatments, conditions and outcomes *prior to statistical analysis*, including those relating to patient identification, therapy, procedures, comorbidities, and components of any composite endpoints.  Preferred: Provided within a pre-published protocol or open-access document *prior to statistical analysis.* | (a) Detailed lists of codes used for each aspect of the study.  (b) Date of publication and access details for the coding manual (please add to box below). (c) Provide definitions, implementation logic and validation of any phenotyping algorithms used. (d) Specify any processes used to validate the coding scheme or reference to prior work. | Select one option:  Minimum standard not met ☐ Minimum standard met ☒  Preferred standard met ☐ |
| 4. Analysis | To fully detail how outcome events were analysed and allow independent assessment of the authenticity of study findings. | Minimum: Describe the process used to analyse study outcomes, including statistical methods and use of any machine learning or algorithmic approaches.  Preferred: Provide a statistical analysis plan as a supplementary file, locked *prior to* *analyses commencing.* | (a) Provide details on all statistical methods used. (b) Provide links to any machine code or algorithms used in the analysis, preferably as open-source. (c) Specify the processes of testing assumptions, assessing model fit and any internal validation.  (d) Specify how generalisability of results was assessed, the replication of findings in other datasets, or any external validation. | Select one option:  Minimum standard not met ☐ Minimum standard met ☒  Preferred standard met ☐ |
| 5. Ethics and governance | To provide patients, who may or may not have given consent, and regulatory authorities the ability to interrogate the security and provenance of the data. | Minimum: Clear unambiguous statements on how the principles of Good Clinical Practice and Data Protection will be/were met, provided in the methods section of the research report.  Preferred: Provided within a pre-published protocol or open-access document with evidence of patient and public engagement. | (a) State how informed consent was acquired, or governance if no patient consent. (b) Specify how data privacy was protected in the collection and storage of data.  (c) Detail what steps were taken for patient and public involvement in the research study. (d) Provide information on where anonymised source data or code can be obtained for verification and further research. | Select one option:  Minimum standard not met ☐ Minimum standard met ☐  Preferred standard met ☒ |
| 6. Coding manual | DOI of publication or website address: https://doi.org/10.21252/t55f-vr93  Date published: 18/Jul/25 | | | |
| 7. Comments | The approved CPRD protocol cannot be made publicly available, but was made available to reviewers of this manuscript as per CPRD guidelines. | | | |
| **8. Summary declaration** | One or more minimum standards not met ☐ OR **All minimum standards met** ☒  Number of preferred standards met: 1 / 5 | | | |

Directions for use:

**Research team:** To complete the checklist, authors will need to consider these points during the design of the research to ensure that coding protocols and coding manuals are pre-published. Where applicable, it is advisable that all five minimum standards are met for an individual research study, whether observational or a controlled trial. If any component is not applicable to the study, the corresponding author can indicate why this is the case in the comment box.

This checklist can accompany the article as a supplementary file on submission to the journal, with the ability for readers to review responses. A comment on the meeting of standards in the text of the method section is suggested, for example; *“this study meets all five of the CODE-EHR minimum framework standards for the use of structured healthcare data in clinical research, with two out of five standards meeting preferred criteria <add reference to this CODE-EHR paper;* [*https://doi.org/10.1136/bmj-2021-069048*](https://doi.org/10.1136/bmj-2021-069048)*>”; OR “this study meets four out of five of the CODE-EHR minimum framework standards for the use of structured healthcare data in clinical research; one of the five minimum standards was not met as coding schemes were not specified prior to analysis <add reference to this CODE-EHR paper;* [*https://doi.org/10.1136/bmj-2021-069048*](https://doi.org/10.1136/bmj-2021-069048)*>.”* Note, easy to complete form versions of this checklist are available in the article appendices (word and pdf versions) and at <https://www.escardio.org/bigdata>.

**Research appraisers (patients, clinicians, regulators, guideline task forces):** Where applicable, it is advisable that all five minimum standards are met for the research study to be considered robust.

**FURTHER DETAILS ON THE CODE-EHR FRAMEWORK:** please refer to Kotecha D, Asselbergs FW, et al; on behalf of the Innovative Medicines Initiative BigData@Heart Consortium, European Society of Cardiology, CODE-EHR international consensus group. CODE-EHR best practice framework for the use of structured electronic healthcare records in clinical research. BMJ 2022;378:e069048. doi:10.1136/bmj-2021-069048. Also published in Lancet Digit Health and Eur Heart J.

Table S2	Prevalence of A&G by Sociodemographic Factors and Year, %

|  |  | 2015 | 2016 | 2017 | 2018 | 2019 | 2020 | 2021 | 2022 | 2023 |
| --- | --- | --- | --- | --- | --- | --- | --- | --- | --- | --- |
| Total |  | 0.10 (0.10 to 0.11) | 0.13 (0.12 to 0.13) | 0.17 (0.17 to 0.17) | 0.34 (0.34 to 0.34) | 0.43 (0.43 to 0.44) | 0.89 (0.88 to 0.90) | 1.31 (1.30 to 1.31) | 1.56 (1.55 to 1.56) | 1.97 (1.96 to 1.98) |
| Gender | Females | 0.08 (0.08 to 0.09) | 0.10 (0.10 to 0.10) | 0.14 (0.14 to 0.14) | 0.28 (0.28 to 0.28) | 0.35 (0.35 to 0.36) | 0.72 (0.71 to 0.73) | 1.03 (1.02 to 1.04) | 1.24 (1.23 to 1.25) | 1.56 (1.55 to 1.57) |
|  | Males | 0.13 (0.12 to 0.13) | 0.15 (0.15 to 0.16) | 0.20 (0.20 to 0.21) | 0.40 (0.40 to 0.41) | 0.52 (0.51 to 0.52) | 1.07 (1.06 to 1.08) | 1.59 (1.58 to 1.60) | 1.88 (1.87 to 1.90) | 2.40 (2.38 to 2.41) |
| Age, years | 0 to 4 | 0.07 (0.06 to 0.08) | 0.07 (0.06 to 0.08) | 0.09 (0.08 to 0.11) | 0.17 (0.16 to 0.19) | 0.22 (0.20 to 0.24) | 0.50 (0.47 to 0.53) | 0.80 (0.76 to 0.83) | 1.06 (1.02 to 1.11) | 1.22 (1.17 to 1.27) |
|  | 5 to 9 | 0.08 (0.07 to 0.08) | 0.06 (0.06 to 0.07) | 0.08 (0.08 to 0.09) | 0.14 (0.14 to 0.15) | 0.19 (0.18 to 0.20) | 0.43 (0.41 to 0.44) | 0.68 (0.66 to 0.70) | 0.83 (0.80 to 0.85) | 1.03 (1.01 to 1.06) |
|  | 10 to 14 | 0.06 (0.06 to 0.07) | 0.07 (0.06 to 0.08) | 0.07 (0.07 to 0.08) | 0.14 (0.13 to 0.15) | 0.18 (0.17 to 0.19) | 0.40 (0.39 to 0.42) | 0.67 (0.65 to 0.69) | 0.78 (0.76 to 0.80) | 0.93 (0.91 to 0.96) |
|  | 15 to 24 | 0.05 (0.05 to 0.06) | 0.06 (0.05 to 0.06) | 0.09 (0.08 to 0.09) | 0.16 (0.16 to 0.17) | 0.20 (0.19 to 0.21) | 0.46 (0.45 to 0.47) | 0.74 (0.73 to 0.76) | 0.83 (0.81 to 0.85) | 1.02 (1.01 to 1.04) |
|  | 25 to 34 | 0.06 (0.06 to 0.07) | 0.08 (0.08 to 0.09) | 0.11 (0.10 to 0.11) | 0.20 (0.19 to 0.20) | 0.25 (0.24 to 0.26) | 0.59 (0.58 to 0.61) | 0.85 (0.84 to 0.87) | 0.94 (0.92 to 0.96) | 1.18 (1.17 to 1.20) |
|  | 35 to 44 | 0.09 (0.08 to 0.09) | 0.10 (0.10 to 0.11) | 0.13 (0.13 to 0.14) | 0.26 (0.25 to 0.27) | 0.33 (0.32 to 0.34) | 0.72 (0.70 to 0.73) | 1.07 (1.05 to 1.09) | 1.22 (1.20 to 1.24) | 1.52 (1.50 to 1.54) |
|  | 45 to 54 | 0.10 (0.10 to 0.11) | 0.13 (0.12 to 0.14) | 0.18 (0.17 to 0.18) | 0.35 (0.34 to 0.36) | 0.44 (0.43 to 0.45) | 0.91 (0.89 to 0.92) | 1.34 (1.32 to 1.36) | 1.61 (1.59 to 1.63) | 2.08 (2.06 to 2.11) |
|  | 55 to 64 | 0.12 (0.12 to 0.13) | 0.15 (0.14 to 0.16) | 0.21 (0.21 to 0.22) | 0.44 (0.43 to 0.45) | 0.54 (0.53 to 0.55) | 1.09 (1.07 to 1.11) | 1.59 (1.57 to 1.61) | 1.94 (1.92 to 1.96) | 2.50 (2.48 to 2.53) |
|  | 65 to 74 | 0.16 (0.16 to 0.17) | 0.20 (0.20 to 0.21) | 0.30 (0.29 to 0.31) | 0.60 (0.58 to 0.61) | 0.77 (0.75 to 0.79) | 1.43 (1.41 to 1.45) | 2.03 (2.01 to 2.06) | 2.52 (2.49 to 2.55) | 3.23 (3.19 to 3.26) |
|  | 75 to 84 | 0.21 (0.20 to 0.23) | 0.28 (0.26 to 0.29) | 0.37 (0.36 to 0.39) | 0.77 (0.75 to 0.80) | 1.01 (0.98 to 1.03) | 1.93 (1.89 to 1.96) | 2.73 (2.69 to 2.76) | 3.29 (3.24 to 3.33) | 4.12 (4.07 to 4.17) |
|  | 85+ | 0.20 (0.19 to 0.22) | 0.27 (0.25 to 0.29) | 0.34 (0.32 to 0.36) | 0.68 (0.65 to 0.71) | 0.95 (0.92 to 0.99) | 1.93 (1.88 to 1.98) | 2.72 (2.66 to 2.78) | 3.37 (3.31 to 3.44) | 4.12 (4.05 to 4.20) |
| Females | 0 to 4 | 0.07 (0.06 to 0.09) | 0.07 (0.06 to 0.09) | 0.11 (0.09 to 0.13) | 0.20 (0.17 to 0.22) | 0.24 (0.21 to 0.27) | 0.55 (0.51 to 0.59) | 0.85 (0.80 to 0.90) | 1.15 (1.09 to 1.21) | 1.34 (1.27 to 1.41) |
|  | 5 to 9 | 0.07 (0.07 to 0.08) | 0.07 (0.06 to 0.07) | 0.09 (0.08 to 0.10) | 0.14 (0.13 to 0.15) | 0.19 (0.18 to 0.21) | 0.43 (0.41 to 0.45) | 0.67 (0.64 to 0.70) | 0.82 (0.79 to 0.85) | 1.02 (0.99 to 1.06) |
|  | 10 to 14 | 0.06 (0.06 to 0.07) | 0.07 (0.06 to 0.08) | 0.07 (0.06 to 0.08) | 0.13 (0.12 to 0.14) | 0.18 (0.16 to 0.19) | 0.37 (0.35 to 0.39) | 0.61 (0.58 to 0.63) | 0.73 (0.71 to 0.76) | 0.90 (0.87 to 0.93) |
|  | 15 to 24 | 0.03 (0.03 to 0.04) | 0.04 (0.04 to 0.05) | 0.06 (0.06 to 0.07) | 0.12 (0.11 to 0.13) | 0.14 (0.13 to 0.15) | 0.31 (0.30 to 0.32) | 0.51 (0.49 to 0.52) | 0.59 (0.57 to 0.61) | 0.72 (0.70 to 0.75) |
|  | 25 to 34 | 0.04 (0.03 to 0.04) | 0.05 (0.04 to 0.05) | 0.07 (0.06 to 0.07) | 0.12 (0.11 to 0.13) | 0.15 (0.14 to 0.16) | 0.34 (0.32 to 0.35) | 0.47 (0.46 to 0.49) | 0.55 (0.53 to 0.56) | 0.66 (0.64 to 0.68) |
|  | 35 to 44 | 0.05 (0.05 to 0.06) | 0.06 (0.06 to 0.07) | 0.09 (0.08 to 0.10) | 0.16 (0.15 to 0.17) | 0.21 (0.20 to 0.22) | 0.43 (0.42 to 0.45) | 0.62 (0.61 to 0.64) | 0.72 (0.70 to 0.74) | 0.89 (0.87 to 0.91) |
|  | 45 to 54 | 0.07 (0.06 to 0.08) | 0.09 (0.08 to 0.09) | 0.12 (0.11 to 0.13) | 0.24 (0.23 to 0.25) | 0.29 (0.28 to 0.31) | 0.62 (0.60 to 0.63) | 0.88 (0.86 to 0.90) | 1.01 (0.99 to 1.04) | 1.33 (1.30 to 1.36) |
|  | 55 to 64 | 0.11 (0.10 to 0.12) | 0.13 (0.12 to 0.13) | 0.18 (0.17 to 0.19) | 0.39 (0.37 to 0.40) | 0.45 (0.43 to 0.46) | 0.92 (0.89 to 0.94) | 1.31 (1.28 to 1.34) | 1.57 (1.54 to 1.60) | 2.02 (1.98 to 2.05) |
|  | 65 to 74 | 0.16 (0.15 to 0.17) | 0.19 (0.18 to 0.21) | 0.28 (0.27 to 0.30) | 0.57 (0.55 to 0.59) | 0.74 (0.72 to 0.76) | 1.39 (1.36 to 1.43) | 1.93 (1.90 to 1.97) | 2.40 (2.36 to 2.44) | 3.07 (3.02 to 3.12) |
|  | 75 to 84 | 0.21 (0.20 to 0.23) | 0.28 (0.26 to 0.30) | 0.37 (0.35 to 0.39) | 0.78 (0.75 to 0.82) | 1.03 (1.00 to 1.07) | 2.02 (1.97 to 2.07) | 2.79 (2.74 to 2.85) | 3.35 (3.29 to 3.41) | 4.17 (4.10 to 4.24) |
|  | 85+ | 0.21 (0.19 to 0.24) | 0.33 (0.30 to 0.36) | 0.38 (0.35 to 0.42) | 0.79 (0.74 to 0.84) | 1.07 (1.01 to 1.14) | 2.23 (2.15 to 2.32) | 3.10 (3.00 to 3.20) | 3.87 (3.75 to 3.98) | 4.66 (4.54 to 4.79) |
| Males | 0 to 4 | 0.07 (0.05 to 0.08) | 0.07 (0.06 to 0.09) | 0.08 (0.06 to 0.10) | 0.15 (0.13 to 0.17) | 0.19 (0.17 to 0.22) | 0.44 (0.41 to 0.48) | 0.74 (0.69 to 0.80) | 0.98 (0.92 to 1.04) | 1.09 (1.03 to 1.16) |
|  | 5 to 9 | 0.08 (0.07 to 0.09) | 0.06 (0.05 to 0.07) | 0.08 (0.07 to 0.09) | 0.15 (0.13 to 0.16) | 0.19 (0.18 to 0.21) | 0.42 (0.40 to 0.45) | 0.69 (0.66 to 0.72) | 0.84 (0.81 to 0.87) | 1.04 (1.01 to 1.08) |
|  | 10 to 14 | 0.06 (0.05 to 0.07) | 0.07 (0.06 to 0.08) | 0.08 (0.07 to 0.09) | 0.16 (0.14 to 0.17) | 0.18 (0.17 to 0.19) | 0.45 (0.42 to 0.47) | 0.74 (0.71 to 0.77) | 0.82 (0.79 to 0.85) | 0.97 (0.94 to 1.00) |
|  | 15 to 24 | 0.07 (0.06 to 0.08) | 0.08 (0.07 to 0.09) | 0.11 (0.10 to 0.12) | 0.21 (0.20 to 0.23) | 0.27 (0.25 to 0.28) | 0.63 (0.61 to 0.65) | 1.01 (0.98 to 1.04) | 1.10 (1.07 to 1.12) | 1.35 (1.32 to 1.39) |
|  | 25 to 34 | 0.10 (0.09 to 0.10) | 0.12 (0.11 to 0.13) | 0.15 (0.14 to 0.16) | 0.28 (0.27 to 0.30) | 0.36 (0.34 to 0.37) | 0.88 (0.86 to 0.91) | 1.28 (1.26 to 1.31) | 1.38 (1.35 to 1.41) | 1.78 (1.75 to 1.81) |
|  | 35 to 44 | 0.12 (0.12 to 0.13) | 0.14 (0.13 to 0.15) | 0.18 (0.17 to 0.19) | 0.36 (0.35 to 0.37) | 0.47 (0.45 to 0.48) | 1.03 (1.01 to 1.05) | 1.55 (1.52 to 1.58) | 1.75 (1.72 to 1.78) | 2.20 (2.16 to 2.23) |
|  | 45 to 54 | 0.13 (0.13 to 0.14) | 0.18 (0.17 to 0.19) | 0.23 (0.22 to 0.24) | 0.46 (0.44 to 0.47) | 0.60 (0.58 to 0.61) | 1.22 (1.19 to 1.24) | 1.84 (1.81 to 1.87) | 2.24 (2.21 to 2.28) | 2.90 (2.86 to 2.94) |
|  | 55 to 64 | 0.14 (0.13 to 0.15) | 0.18 (0.17 to 0.19) | 0.24 (0.23 to 0.26) | 0.50 (0.48 to 0.51) | 0.64 (0.62 to 0.66) | 1.27 (1.24 to 1.29) | 1.88 (1.85 to 1.92) | 2.32 (2.29 to 2.36) | 3.01 (2.97 to 3.05) |
|  | 65 to 74 | 0.17 (0.16 to 0.18) | 0.21 (0.20 to 0.23) | 0.31 (0.30 to 0.33) | 0.62 (0.60 to 0.64) | 0.80 (0.78 to 0.83) | 1.47 (1.43 to 1.50) | 2.13 (2.09 to 2.17) | 2.64 (2.59 to 2.68) | 3.38 (3.33 to 3.43) |
|  | 75 to 84 | 0.22 (0.20 to 0.23) | 0.27 (0.26 to 0.29) | 0.38 (0.36 to 0.40) | 0.77 (0.74 to 0.79) | 0.98 (0.95 to 1.01) | 1.84 (1.80 to 1.89) | 2.67 (2.61 to 2.72) | 3.23 (3.18 to 3.29) | 4.07 (4.01 to 4.14) |
|  | 85+ | 0.20 (0.18 to 0.22) | 0.23 (0.21 to 0.25) | 0.32 (0.29 to 0.34) | 0.61 (0.57 to 0.64) | 0.88 (0.84 to 0.93) | 1.74 (1.68 to 1.80) | 2.48 (2.41 to 2.55) | 3.06 (2.98 to 3.14) | 3.77 (3.68 to 3.86) |
| Ethnicity | Asian | 0.07 (0.07 to 0.08) | 0.08 (0.08 to 0.09) | 0.10 (0.10 to 0.11) | 0.19 (0.18 to 0.20) | 0.25 (0.24 to 0.26) | 0.84 (0.82 to 0.85) | 1.33 (1.31 to 1.35) | 1.51 (1.49 to 1.53) | 1.85 (1.82 to 1.87) |
|  | Black | 0.08 (0.07 to 0.09) | 0.09 (0.08 to 0.10) | 0.11 (0.10 to 0.12) | 0.21 (0.20 to 0.23) | 0.32 (0.31 to 0.34) | 0.83 (0.80 to 0.85) | 1.20 (1.17 to 1.23) | 1.39 (1.35 to 1.42) | 1.76 (1.72 to 1.79) |
|  | Mixed/Multiple | 0.06 (0.05 to 0.08) | 0.08 (0.06 to 0.09) | 0.08 (0.07 to 0.10) | 0.16 (0.14 to 0.18) | 0.23 (0.21 to 0.26) | 0.61 (0.58 to 0.64) | 0.96 (0.92 to 1.00) | 1.13 (1.09 to 1.17) | 1.42 (1.37 to 1.47) |
|  | White | 0.11 (0.11 to 0.12) | 0.14 (0.14 to 0.14) | 0.19 (0.19 to 0.19) | 0.38 (0.38 to 0.39) | 0.48 (0.48 to 0.49) | 0.94 (0.94 to 0.95) | 1.37 (1.36 to 1.38) | 1.64 (1.64 to 1.65) | 2.10 (2.09 to 2.11) |
| IMD Quintile | Least | 0.09 (0.08 to 0.09) | 0.13 (0.12 to 0.13) | 0.17 (0.16 to 0.17) | 0.33 (0.33 to 0.34) | 0.43 (0.42 to 0.44) | 0.91 (0.90 to 0.92) | 1.36 (1.34 to 1.37) | 1.67 (1.65 to 1.69) | 2.15 (2.13 to 2.17) |
|  | Second Least | 0.12 (0.12 to 0.13) | 0.15 (0.15 to 0.16) | 0.20 (0.19 to 0.20) | 0.38 (0.37 to 0.38) | 0.49 (0.48 to 0.50) | 0.94 (0.93 to 0.95) | 1.37 (1.35 to 1.38) | 1.66 (1.64 to 1.67) | 2.07 (2.05 to 2.09) |
|  | Neither | 0.12 (0.11 to 0.12) | 0.14 (0.14 to 0.15) | 0.20 (0.19 to 0.20) | 0.39 (0.38 to 0.40) | 0.50 (0.49 to 0.51) | 0.97 (0.96 to 0.99) | 1.42 (1.41 to 1.44) | 1.65 (1.63 to 1.67) | 2.04 (2.02 to 2.06) |
|  | Second Most | 0.11 (0.10 to 0.11) | 0.12 (0.12 to 0.13) | 0.16 (0.15 to 0.16) | 0.33 (0.32 to 0.34) | 0.41 (0.40 to 0.42) | 0.89 (0.88 to 0.90) | 1.31 (1.30 to 1.33) | 1.53 (1.51 to 1.54) | 1.88 (1.86 to 1.89) |
|  | Most | 0.09 (0.08 to 0.09) | 0.09 (0.09 to 0.10) | 0.15 (0.14 to 0.15) | 0.27 (0.27 to 0.28) | 0.35 (0.34 to 0.35) | 0.74 (0.73 to 0.76) | 1.08 (1.06 to 1.09) | 1.29 (1.28 to 1.31) | 1.75 (1.73 to 1.77) |
| Asian | Least | 0.07 (0.05 to 0.09) | 0.09 (0.07 to 0.11) | 0.14 (0.11 to 0.16) | 0.24 (0.21 to 0.27) | 0.24 (0.21 to 0.27) | 0.91 (0.86 to 0.97) | 1.46 (1.39 to 1.53) | 1.68 (1.61 to 1.76) | 2.00 (1.92 to 2.08) |
|  | Second Least | 0.07 (0.05 to 0.09) | 0.09 (0.07 to 0.11) | 0.10 (0.08 to 0.12) | 0.16 (0.14 to 0.19) | 0.21 (0.19 to 0.24) | 0.86 (0.81 to 0.91) | 1.35 (1.29 to 1.41) | 1.51 (1.45 to 1.57) | 1.72 (1.66 to 1.79) |
|  | Neither | 0.08 (0.07 to 0.10) | 0.10 (0.08 to 0.11) | 0.11 (0.09 to 0.12) | 0.18 (0.16 to 0.20) | 0.25 (0.23 to 0.27) | 1.02 (0.97 to 1.06) | 1.58 (1.53 to 1.64) | 1.75 (1.69 to 1.80) | 1.91 (1.86 to 1.97) |
|  | Second Most | 0.09 (0.07 to 0.10) | 0.07 (0.06 to 0.09) | 0.08 (0.07 to 0.09) | 0.20 (0.18 to 0.21) | 0.28 (0.26 to 0.30) | 0.90 (0.87 to 0.94) | 1.41 (1.37 to 1.46) | 1.56 (1.52 to 1.61) | 1.87 (1.83 to 1.92) |
|  | Most | 0.06 (0.05 to 0.07) | 0.08 (0.07 to 0.09) | 0.11 (0.10 to 0.12) | 0.18 (0.16 to 0.20) | 0.24 (0.22 to 0.26) | 0.58 (0.55 to 0.61) | 0.98 (0.95 to 1.02) | 1.20 (1.16 to 1.24) | 1.76 (1.72 to 1.81) |
| Black | Least | 0.05 (0.02 to 0.10) | 0.06 (0.03 to 0.12) | 0.16 (0.11 to 0.24) | 0.27 (0.20 to 0.35) | 0.26 (0.20 to 0.35) | 0.84 (0.73 to 0.98) | 1.14 (1.00 to 1.29) | 1.61 (1.45 to 1.79) | 2.06 (1.88 to 2.26) |
|  | Second Least | 0.07 (0.05 to 0.11) | 0.11 (0.08 to 0.16) | 0.14 (0.11 to 0.19) | 0.21 (0.16 to 0.27) | 0.31 (0.25 to 0.38) | 0.93 (0.83 to 1.04) | 1.27 (1.16 to 1.40) | 1.49 (1.37 to 1.63) | 1.92 (1.78 to 2.07) |
|  | Neither | 0.08 (0.06 to 0.10) | 0.09 (0.07 to 0.12) | 0.11 (0.09 to 0.14) | 0.21 (0.18 to 0.25) | 0.27 (0.23 to 0.31) | 0.86 (0.80 to 0.93) | 1.28 (1.20 to 1.36) | 1.41 (1.33 to 1.50) | 1.76 (1.67 to 1.86) |
|  | Second Most | 0.09 (0.07 to 0.10) | 0.09 (0.08 to 0.11) | 0.09 (0.08 to 0.11) | 0.24 (0.21 to 0.26) | 0.36 (0.34 to 0.39) | 0.91 (0.87 to 0.96) | 1.30 (1.24 to 1.35) | 1.50 (1.45 to 1.56) | 1.81 (1.75 to 1.87) |
|  | Most | 0.07 (0.06 to 0.09) | 0.09 (0.07 to 0.10) | 0.11 (0.09 to 0.12) | 0.19 (0.17 to 0.21) | 0.32 (0.29 to 0.35) | 0.72 (0.68 to 0.76) | 1.07 (1.03 to 1.12) | 1.23 (1.19 to 1.28) | 1.65 (1.60 to 1.71) |
| Mixed/Multiple | Least | 0.03 (0.01 to 0.06) | 0.07 (0.04 to 0.11) | 0.10 (0.07 to 0.15) | 0.16 (0.12 to 0.22) | 0.18 (0.14 to 0.24) | 0.56 (0.48 to 0.65) | 0.95 (0.85 to 1.06) | 1.33 (1.21 to 1.46) | 1.45 (1.33 to 1.59) |
|  | Second Least | 0.08 (0.05 to 0.13) | 0.09 (0.06 to 0.14) | 0.11 (0.08 to 0.16) | 0.18 (0.14 to 0.24) | 0.23 (0.18 to 0.29) | 0.49 (0.42 to 0.57) | 1.03 (0.93 to 1.14) | 1.18 (1.07 to 1.30) | 1.41 (1.29 to 1.54) |
|  | Neither | 0.08 (0.05 to 0.12) | 0.11 (0.08 to 0.15) | 0.08 (0.05 to 0.12) | 0.18 (0.14 to 0.24) | 0.25 (0.21 to 0.31) | 0.67 (0.59 to 0.75) | 1.08 (0.99 to 1.19) | 1.13 (1.03 to 1.24) | 1.45 (1.34 to 1.57) |
|  | Second Most | 0.07 (0.05 to 0.10) | 0.07 (0.05 to 0.11) | 0.07 (0.05 to 0.10) | 0.15 (0.12 to 0.19) | 0.25 (0.21 to 0.30) | 0.71 (0.65 to 0.79) | 0.96 (0.88 to 1.04) | 1.19 (1.10 to 1.28) | 1.46 (1.37 to 1.56) |
|  | Most | 0.05 (0.04 to 0.08) | 0.06 (0.04 to 0.08) | 0.08 (0.06 to 0.11) | 0.15 (0.12 to 0.18) | 0.24 (0.20 to 0.28) | 0.57 (0.51 to 0.64) | 0.83 (0.76 to 0.91) | 0.94 (0.86 to 1.02) | 1.34 (1.25 to 1.44) |
| White | Least | 0.09 (0.09 to 0.10) | 0.14 (0.13 to 0.14) | 0.18 (0.17 to 0.18) | 0.35 (0.35 to 0.36) | 0.47 (0.46 to 0.48) | 0.95 (0.94 to 0.97) | 1.41 (1.39 to 1.43) | 1.73 (1.71 to 1.75) | 2.24 (2.22 to 2.26) |
|  | Second Least | 0.13 (0.13 to 0.14) | 0.16 (0.16 to 0.17) | 0.21 (0.21 to 0.22) | 0.41 (0.40 to 0.42) | 0.53 (0.52 to 0.55) | 0.99 (0.97 to 1.00) | 1.43 (1.41 to 1.44) | 1.75 (1.73 to 1.76) | 2.20 (2.17 to 2.22) |
|  | Neither | 0.13 (0.13 to 0.14) | 0.15 (0.15 to 0.16) | 0.22 (0.21 to 0.23) | 0.44 (0.43 to 0.45) | 0.57 (0.55 to 0.58) | 1.02 (1.00 to 1.04) | 1.47 (1.45 to 1.49) | 1.73 (1.71 to 1.75) | 2.17 (2.15 to 2.19) |
|  | Second Most | 0.12 (0.11 to 0.13) | 0.14 (0.13 to 0.14) | 0.18 (0.18 to 0.19) | 0.39 (0.38 to 0.40) | 0.46 (0.45 to 0.47) | 0.93 (0.91 to 0.94) | 1.37 (1.35 to 1.39) | 1.60 (1.58 to 1.62) | 1.98 (1.96 to 2.01) |
|  | Most | 0.10 (0.09 to 0.10) | 0.10 (0.09 to 0.10) | 0.17 (0.16 to 0.17) | 0.32 (0.31 to 0.33) | 0.39 (0.38 to 0.40) | 0.81 (0.80 to 0.83) | 1.14 (1.13 to 1.16) | 1.38 (1.37 to 1.40) | 1.86 (1.84 to 1.88) |
| English Region | London | 0.09 (0.09 to 0.10) | 0.10 (0.09 to 0.10) | 0.09 (0.08 to 0.09) | 0.19 (0.19 to 0.20) | 0.34 (0.33 to 0.35) | 1.09 (1.07 to 1.10) | 1.59 (1.57 to 1.60) | 1.81 (1.79 to 1.83) | 2.16 (2.14 to 2.17) |
|  | Midlands/East | 0.11 (0.11 to 0.12) | 0.19 (0.18 to 0.19) | 0.22 (0.22 to 0.23) | 0.39 (0.38 to 0.39) | 0.46 (0.45 to 0.47) | 0.79 (0.78 to 0.80) | 1.29 (1.27 to 1.30) | 1.69 (1.67 to 1.70) | 2.09 (2.07 to 2.10) |
|  | North England | 0.04 (0.04 to 0.05) | 0.04 (0.04 to 0.04) | 0.09 (0.08 to 0.09) | 0.27 (0.26 to 0.27) | 0.31 (0.31 to 0.32) | 0.66 (0.65 to 0.67) | 0.87 (0.85 to 0.88) | 1.06 (1.04 to 1.07) | 1.32 (1.30 to 1.33) |
|  | South England | 0.16 (0.15 to 0.16) | 0.17 (0.17 to 0.18) | 0.25 (0.25 to 0.26) | 0.46 (0.45 to 0.46) | 0.57 (0.56 to 0.58) | 1.02 (1.01 to 1.03) | 1.48 (1.47 to 1.50) | 1.69 (1.67 to 1.70) | 2.29 (2.27 to 2.30) |

Table S3	Prevalence of Direct Referrals by Sociodemographic Factors and Year, %

|  |  | 2015 | 2016 | 2017 | 2018 | 2019 | 2020 | 2021 | 2022 | 2023 |
| --- | --- | --- | --- | --- | --- | --- | --- | --- | --- | --- |
| Total |  | 24.6 (24.6 to 24.6) | 24.1 (24.0 to 24.1) | 23.1 (23.1 to 23.2) | 23.4 (23.4 to 23.5) | 23.8 (23.8 to 23.8) | 17.8 (17.7 to 17.8) | 21.1 (21.1 to 21.1) | 22.8 (22.8 to 22.8) | 23.6 (23.6 to 23.6) |
| Gender | Females | 21.0 (20.9 to 21.0) | 20.5 (20.5 to 20.6) | 19.7 (19.7 to 19.8) | 20.0 (19.9 to 20.0) | 20.2 (20.1 to 20.2) | 14.7 (14.7 to 14.8) | 17.4 (17.4 to 17.4) | 19.1 (19.0 to 19.1) | 19.7 (19.7 to 19.7) |
|  | Males | 28.3 (28.3 to 28.4) | 27.7 (27.7 to 27.8) | 26.7 (26.6 to 26.7) | 27.0 (27.0 to 27.1) | 27.6 (27.5 to 27.6) | 20.9 (20.9 to 21.0) | 24.9 (24.9 to 25.0) | 26.6 (26.6 to 26.7) | 27.6 (27.6 to 27.7) |
| Age, years | 0 to 4 | 13.5 (13.3 to 13.6) | 13.3 (13.1 to 13.4) | 12.3 (12.2 to 12.4) | 12.4 (12.2 to 12.5) | 12.4 (12.3 to 12.6) | 8.7 (8.5 to 8.8) | 11.3 (11.2 to 11.4) | 13.5 (13.4 to 13.7) | 13.4 (13.3 to 13.6) |
|  | 5 to 9 | 11.4 (11.3 to 11.5) | 11.7 (11.6 to 11.7) | 11.3 (11.3 to 11.4) | 11.4 (11.3 to 11.5) | 11.4 (11.3 to 11.5) | 8.0 (8.0 to 8.1) | 9.9 (9.8 to 10.0) | 11.7 (11.6 to 11.8) | 12.5 (12.4 to 12.6) |
|  | 10 to 14 | 11.2 (11.1 to 11.3) | 11.5 (11.4 to 11.6) | 11.3 (11.2 to 11.4) | 11.4 (11.3 to 11.5) | 11.6 (11.5 to 11.7) | 8.2 (8.2 to 8.3) | 10.6 (10.6 to 10.7) | 11.5 (11.5 to 11.6) | 11.9 (11.8 to 12.0) |
|  | 15 to 24 | 14.2 (14.1 to 14.3) | 13.8 (13.7 to 13.9) | 13.2 (13.2 to 13.3) | 13.3 (13.2 to 13.4) | 13.4 (13.3 to 13.5) | 10.1 (10.1 to 10.2) | 12.6 (12.5 to 12.6) | 13.1 (13.0 to 13.2) | 13.5 (13.4 to 13.5) |
|  | 25 to 34 | 17.7 (17.6 to 17.8) | 17.2 (17.1 to 17.3) | 16.3 (16.2 to 16.4) | 16.0 (16.0 to 16.1) | 16.0 (15.9 to 16.0) | 12.2 (12.2 to 12.3) | 14.4 (14.3 to 14.4) | 14.9 (14.8 to 15.0) | 15.4 (15.4 to 15.5) |
|  | 35 to 44 | 22.1 (22.0 to 22.2) | 21.4 (21.4 to 21.5) | 20.6 (20.5 to 20.6) | 20.5 (20.4 to 20.6) | 20.6 (20.5 to 20.7) | 15.3 (15.2 to 15.4) | 18.1 (18.1 to 18.2) | 19.2 (19.1 to 19.2) | 19.8 (19.7 to 19.8) |
|  | 45 to 54 | 26.3 (26.2 to 26.4) | 25.7 (25.6 to 25.8) | 24.8 (24.7 to 24.9) | 25.1 (25.0 to 25.2) | 25.8 (25.7 to 25.8) | 19.0 (19.0 to 19.1) | 22.7 (22.6 to 22.8) | 24.3 (24.2 to 24.4) | 25.1 (25.0 to 25.1) |
|  | 55 to 64 | 30.4 (30.3 to 30.5) | 29.8 (29.7 to 29.9) | 28.6 (28.6 to 28.7) | 29.3 (29.2 to 29.4) | 30.0 (30.0 to 30.1) | 22.2 (22.1 to 22.3) | 26.3 (26.2 to 26.4) | 28.9 (28.8 to 29.0) | 30.0 (30.0 to 30.1) |
|  | 65 to 74 | 36.6 (36.5 to 36.8) | 35.9 (35.8 to 36.0) | 34.6 (34.5 to 34.7) | 35.6 (35.4 to 35.7) | 36.6 (36.5 to 36.7) | 26.9 (26.8 to 27.0) | 31.6 (31.5 to 31.7) | 34.8 (34.7 to 34.9) | 36.0 (35.9 to 36.1) |
|  | 75 to 84 | 45.2 (45.0 to 45.4) | 44.3 (44.1 to 44.5) | 42.8 (42.7 to 43.0) | 44.2 (44.0 to 44.3) | 45.1 (45.0 to 45.3) | 34.4 (34.3 to 34.6) | 40.2 (40.1 to 40.4) | 43.5 (43.3 to 43.6) | 44.5 (44.3 to 44.6) |
|  | 85+ | 46.0 (45.7 to 46.2) | 46.1 (45.9 to 46.4) | 44.5 (44.2 to 44.7) | 46.0 (45.8 to 46.3) | 47.2 (46.9 to 47.4) | 38.6 (38.4 to 38.9) | 44.9 (44.7 to 45.2) | 47.9 (47.6 to 48.1) | 48.5 (48.3 to 48.8) |
| Females | 0 to 4 | 14.9 (14.7 to 15.1) | 14.6 (14.4 to 14.8) | 13.7 (13.5 to 13.9) | 13.6 (13.4 to 13.9) | 13.8 (13.6 to 14.0) | 9.6 (9.4 to 9.8) | 12.6 (12.4 to 12.8) | 15.0 (14.8 to 15.3) | 14.9 (14.6 to 15.1) |
|  | 5 to 9 | 12.2 (12.1 to 12.4) | 12.5 (12.4 to 12.6) | 12.2 (12.1 to 12.3) | 12.2 (12.1 to 12.3) | 12.3 (12.1 to 12.4) | 8.5 (8.4 to 8.6) | 10.6 (10.5 to 10.8) | 12.5 (12.3 to 12.6) | 13.3 (13.2 to 13.4) |
|  | 10 to 14 | 11.0 (10.9 to 11.2) | 11.3 (11.2 to 11.4) | 11.1 (11.0 to 11.2) | 11.2 (11.1 to 11.3) | 11.4 (11.3 to 11.6) | 8.0 (7.9 to 8.1) | 10.2 (10.1 to 10.3) | 11.2 (11.1 to 11.4) | 11.6 (11.5 to 11.7) |
|  | 15 to 24 | 11.1 (11.0 to 11.1) | 10.8 (10.7 to 10.8) | 10.5 (10.4 to 10.6) | 10.6 (10.5 to 10.7) | 10.7 (10.6 to 10.8) | 7.7 (7.6 to 7.8) | 9.5 (9.4 to 9.5) | 10.2 (10.1 to 10.2) | 10.4 (10.3 to 10.5) |
|  | 25 to 34 | 12.0 (11.9 to 12.1) | 11.7 (11.6 to 11.8) | 11.0 (10.9 to 11.1) | 10.9 (10.8 to 11.0) | 10.9 (10.8 to 10.9) | 7.9 (7.8 to 7.9) | 9.3 (9.3 to 9.4) | 10.0 (9.9 to 10.1) | 10.4 (10.3 to 10.4) |
|  | 35 to 44 | 16.5 (16.4 to 16.6) | 15.9 (15.8 to 16.0) | 15.1 (15.1 to 15.2) | 14.9 (14.8 to 15.0) | 14.9 (14.8 to 15.0) | 10.4 (10.4 to 10.5) | 12.3 (12.2 to 12.4) | 13.4 (13.3 to 13.5) | 13.8 (13.7 to 13.9) |
|  | 45 to 54 | 21.3 (21.2 to 21.4) | 20.7 (20.6 to 20.8) | 19.8 (19.7 to 19.9) | 19.9 (19.8 to 20.0) | 20.1 (20.0 to 20.2) | 14.3 (14.2 to 14.4) | 16.8 (16.7 to 16.9) | 18.3 (18.2 to 18.4) | 18.8 (18.7 to 18.9) |
|  | 55 to 64 | 27.5 (27.4 to 27.6) | 26.8 (26.7 to 26.9) | 25.7 (25.6 to 25.8) | 26.2 (26.1 to 26.3) | 26.4 (26.3 to 26.5) | 19.3 (19.2 to 19.4) | 22.6 (22.5 to 22.7) | 25.0 (24.9 to 25.1) | 25.8 (25.7 to 26.0) |
|  | 65 to 74 | 35.2 (35.0 to 35.4) | 34.5 (34.3 to 34.6) | 33.2 (33.1 to 33.4) | 34.3 (34.2 to 34.5) | 35.1 (34.9 to 35.2) | 26.0 (25.8 to 26.1) | 30.1 (30.0 to 30.3) | 33.4 (33.2 to 33.5) | 34.4 (34.2 to 34.5) |
|  | 75 to 84 | 44.7 (44.5 to 45.0) | 43.8 (43.6 to 44.1) | 42.3 (42.1 to 42.5) | 43.7 (43.5 to 43.9) | 44.5 (44.2 to 44.7) | 34.2 (34.0 to 34.4) | 39.6 (39.4 to 39.8) | 42.9 (42.7 to 43.1) | 43.9 (43.7 to 44.1) |
|  | 85+ | 47.3 (46.9 to 47.8) | 47.8 (47.4 to 48.2) | 46.0 (45.6 to 46.4) | 47.9 (47.5 to 48.3) | 48.9 (48.5 to 49.3) | 40.4 (40.0 to 40.8) | 46.5 (46.1 to 46.9) | 49.6 (49.2 to 50.0) | 50.4 (50.0 to 50.8) |
| Males | 0 to 4 | 12.0 (11.8 to 12.2) | 11.9 (11.7 to 12.1) | 10.8 (10.6 to 11.0) | 11.0 (10.8 to 11.2) | 11.0 (10.8 to 11.2) | 7.7 (7.6 to 7.9) | 9.9 (9.8 to 10.1) | 11.9 (11.7 to 12.2) | 11.9 (11.7 to 12.1) |
|  | 5 to 9 | 10.6 (10.5 to 10.7) | 10.8 (10.7 to 10.9) | 10.5 (10.3 to 10.6) | 10.5 (10.4 to 10.7) | 10.5 (10.4 to 10.6) | 7.5 (7.4 to 7.6) | 9.1 (9.0 to 9.2) | 10.9 (10.8 to 11.1) | 11.7 (11.6 to 11.8) |
|  | 10 to 14 | 11.4 (11.3 to 11.5) | 11.6 (11.5 to 11.8) | 11.5 (11.3 to 11.6) | 11.6 (11.5 to 11.7) | 11.7 (11.6 to 11.8) | 8.5 (8.5 to 8.6) | 11.1 (11.0 to 11.3) | 11.8 (11.7 to 11.9) | 12.2 (12.1 to 12.4) |
|  | 15 to 24 | 18.0 (17.9 to 18.1) | 17.4 (17.3 to 17.5) | 16.5 (16.4 to 16.6) | 16.5 (16.4 to 16.6) | 16.5 (16.4 to 16.6) | 12.9 (12.8 to 13.0) | 16.0 (15.9 to 16.1) | 16.4 (16.3 to 16.5) | 16.8 (16.7 to 16.9) |
|  | 25 to 34 | 23.9 (23.8 to 24.1) | 23.3 (23.2 to 23.4) | 22.1 (22.0 to 22.2) | 21.7 (21.6 to 21.8) | 21.7 (21.6 to 21.8) | 17.1 (17.0 to 17.2) | 20.0 (19.9 to 20.1) | 20.4 (20.3 to 20.5) | 21.2 (21.1 to 21.3) |
|  | 35 to 44 | 28.1 (27.9 to 28.2) | 27.4 (27.2 to 27.5) | 26.3 (26.2 to 26.5) | 26.5 (26.3 to 26.6) | 26.7 (26.6 to 26.9) | 20.6 (20.5 to 20.7) | 24.4 (24.3 to 24.5) | 25.3 (25.2 to 25.4) | 26.2 (26.1 to 26.3) |
|  | 45 to 54 | 31.6 (31.5 to 31.7) | 31.0 (30.9 to 31.2) | 30.0 (29.9 to 30.1) | 30.6 (30.5 to 30.8) | 31.8 (31.7 to 31.9) | 24.1 (24.0 to 24.2) | 29.0 (28.9 to 29.1) | 30.7 (30.6 to 30.9) | 31.8 (31.7 to 31.9) |
|  | 55 to 64 | 33.4 (33.2 to 33.5) | 32.8 (32.7 to 33.0) | 31.7 (31.6 to 31.9) | 32.6 (32.4 to 32.7) | 33.9 (33.7 to 34.0) | 25.3 (25.1 to 25.4) | 30.1 (30.0 to 30.2) | 33.0 (32.8 to 33.1) | 34.4 (34.3 to 34.6) |
|  | 65 to 74 | 38.0 (37.8 to 38.2) | 37.3 (37.1 to 37.4) | 35.9 (35.7 to 36.0) | 36.7 (36.6 to 36.9) | 38.1 (37.9 to 38.2) | 27.8 (27.7 to 28.0) | 33.0 (32.8 to 33.1) | 36.2 (36.0 to 36.4) | 37.5 (37.4 to 37.7) |
|  | 75 to 84 | 45.6 (45.4 to 45.8) | 44.7 (44.5 to 45.0) | 43.3 (43.1 to 43.5) | 44.5 (44.3 to 44.8) | 45.7 (45.5 to 45.9) | 34.6 (34.4 to 34.8) | 40.7 (40.5 to 40.9) | 43.9 (43.7 to 44.2) | 45.0 (44.8 to 45.2) |
|  | 85+ | 45.2 (44.9 to 45.5) | 45.2 (44.9 to 45.5) | 43.5 (43.2 to 43.8) | 44.8 (44.5 to 45.2) | 46.1 (45.8 to 46.4) | 37.6 (37.3 to 37.8) | 44.0 (43.7 to 44.3) | 46.7 (46.4 to 47.1) | 47.4 (47.0 to 47.7) |
| Ethnicity | Asian | 20.9 (20.8 to 21.0) | 20.7 (20.6 to 20.8) | 20.6 (20.5 to 20.7) | 20.8 (20.7 to 20.9) | 20.9 (20.8 to 21.0) | 14.8 (14.8 to 14.9) | 18.1 (18.1 to 18.2) | 19.6 (19.5 to 19.7) | 20.4 (20.4 to 20.5) |
|  | Black | 20.9 (20.7 to 21.0) | 20.8 (20.7 to 20.9) | 20.3 (20.2 to 20.4) | 20.8 (20.6 to 20.9) | 21.1 (21.0 to 21.3) | 15.7 (15.5 to 15.8) | 18.9 (18.7 to 19.0) | 20.8 (20.7 to 20.9) | 21.4 (21.2 to 21.5) |
|  | Mixed/Multiple | 18.1 (17.9 to 18.3) | 17.6 (17.4 to 17.8) | 17.0 (16.8 to 17.2) | 17.3 (17.1 to 17.4) | 17.4 (17.2 to 17.6) | 12.9 (12.7 to 13.0) | 15.7 (15.5 to 15.9) | 17.1 (16.9 to 17.3) | 17.6 (17.4 to 17.8) |
|  | White | 26.2 (26.2 to 26.2) | 25.6 (25.6 to 25.6) | 24.5 (24.5 to 24.5) | 24.9 (24.8 to 24.9) | 25.3 (25.3 to 25.3) | 19.0 (19.0 to 19.1) | 22.5 (22.5 to 22.5) | 24.3 (24.2 to 24.3) | 25.1 (25.1 to 25.2) |
| IMD Quintile | Least | 25.6 (25.5 to 25.6) | 25.1 (25.0 to 25.2) | 24.2 (24.1 to 24.2) | 24.6 (24.5 to 24.6) | 25.0 (24.9 to 25.1) | 18.7 (18.6 to 18.7) | 22.1 (22.0 to 22.2) | 24.0 (23.9 to 24.1) | 24.8 (24.8 to 24.9) |
|  | Second Least | 25.3 (25.2 to 25.4) | 24.6 (24.5 to 24.7) | 23.6 (23.6 to 23.7) | 23.9 (23.9 to 24.0) | 24.5 (24.4 to 24.5) | 18.3 (18.2 to 18.3) | 21.6 (21.5 to 21.6) | 23.4 (23.3 to 23.4) | 24.3 (24.2 to 24.3) |
|  | Neither | 24.6 (24.5 to 24.7) | 24.0 (23.9 to 24.1) | 22.8 (22.8 to 22.9) | 23.2 (23.1 to 23.3) | 23.7 (23.6 to 23.7) | 17.7 (17.6 to 17.7) | 20.9 (20.8 to 21.0) | 22.7 (22.6 to 22.7) | 23.5 (23.4 to 23.6) |
|  | Second Most | 23.8 (23.7 to 23.9) | 23.3 (23.3 to 23.4) | 22.5 (22.4 to 22.5) | 22.7 (22.7 to 22.8) | 23.0 (22.9 to 23.1) | 17.2 (17.1 to 17.2) | 20.4 (20.4 to 20.5) | 22.0 (21.9 to 22.0) | 22.7 (22.7 to 22.8) |
|  | Most | 23.9 (23.9 to 24.0) | 23.6 (23.5 to 23.7) | 22.8 (22.7 to 22.8) | 23.0 (22.9 to 23.0) | 23.1 (23.1 to 23.2) | 17.3 (17.3 to 17.4) | 20.8 (20.7 to 20.8) | 22.2 (22.1 to 22.3) | 23.0 (22.9 to 23.1) |
| Asian | Least | 23.8 (23.4 to 24.1) | 23.4 (23.1 to 23.7) | 22.2 (21.9 to 22.6) | 22.1 (21.8 to 22.3) | 22.1 (21.8 to 22.4) | 15.7 (15.4 to 15.9) | 19.2 (19.0 to 19.5) | 20.5 (20.3 to 20.8) | 21.3 (21.0 to 21.5) |
|  | Second Least | 22.0 (21.7 to 22.2) | 21.3 (21.0 to 21.5) | 20.8 (20.6 to 21.1) | 20.8 (20.6 to 21.1) | 21.1 (20.9 to 21.4) | 14.9 (14.7 to 15.1) | 18.0 (17.8 to 18.3) | 19.6 (19.3 to 19.8) | 20.3 (20.1 to 20.6) |
|  | Neither | 21.2 (20.9 to 21.4) | 20.9 (20.7 to 21.1) | 20.6 (20.3 to 20.8) | 20.8 (20.5 to 21.0) | 21.0 (20.7 to 21.2) | 14.9 (14.7 to 15.0) | 18.2 (18.0 to 18.4) | 19.5 (19.4 to 19.7) | 20.4 (20.2 to 20.6) |
|  | Second Most | 19.9 (19.7 to 20.0) | 19.9 (19.7 to 20.1) | 19.9 (19.7 to 20.1) | 20.5 (20.3 to 20.7) | 20.4 (20.2 to 20.5) | 14.5 (14.3 to 14.6) | 17.5 (17.3 to 17.6) | 19.0 (18.8 to 19.2) | 19.7 (19.5 to 19.9) |
|  | Most | 20.3 (20.1 to 20.5) | 20.1 (19.9 to 20.3) | 20.5 (20.3 to 20.7) | 20.5 (20.3 to 20.7) | 20.7 (20.6 to 20.9) | 14.9 (14.8 to 15.1) | 18.4 (18.2 to 18.5) | 20.0 (19.8 to 20.1) | 21.0 (20.9 to 21.2) |
| Black | Least | 22.0 (21.3 to 22.8) | 21.7 (21.0 to 22.5) | 20.9 (20.2 to 21.6) | 21.3 (20.6 to 22.0) | 21.0 (20.4 to 21.7) | 15.8 (15.2 to 16.3) | 18.9 (18.3 to 19.5) | 20.2 (19.6 to 20.8) | 21.1 (20.5 to 21.8) |
|  | Second Least | 22.0 (21.5 to 22.6) | 21.3 (20.8 to 21.9) | 21.1 (20.5 to 21.6) | 21.2 (20.7 to 21.7) | 21.5 (21.0 to 22.0) | 16.0 (15.5 to 16.4) | 19.3 (18.9 to 19.8) | 20.9 (20.4 to 21.4) | 21.4 (20.9 to 21.9) |
|  | Neither | 22.1 (21.7 to 22.4) | 21.6 (21.3 to 22.0) | 21.0 (20.7 to 21.4) | 21.3 (20.9 to 21.6) | 21.7 (21.4 to 22.1) | 16.3 (16.0 to 16.6) | 19.5 (19.1 to 19.8) | 21.5 (21.2 to 21.8) | 22.3 (22.0 to 22.6) |
|  | Second Most | 21.1 (20.9 to 21.4) | 20.9 (20.7 to 21.2) | 20.5 (20.3 to 20.8) | 21.3 (21.0 to 21.5) | 21.8 (21.6 to 22.1) | 16.0 (15.9 to 16.2) | 19.5 (19.3 to 19.7) | 21.5 (21.3 to 21.7) | 22.0 (21.8 to 22.3) |
|  | Most | 19.9 (19.7 to 20.1) | 20.2 (20.0 to 20.4) | 19.7 (19.5 to 19.9) | 20.0 (19.8 to 20.2) | 20.3 (20.1 to 20.5) | 15.0 (14.8 to 15.2) | 18.0 (17.8 to 18.2) | 20.0 (19.8 to 20.2) | 20.5 (20.3 to 20.7) |
| Mixed/Multiple | Least | 18.6 (18.0 to 19.2) | 18.0 (17.5 to 18.6) | 17.5 (17.0 to 18.0) | 17.3 (16.8 to 17.8) | 17.1 (16.6 to 17.5) | 12.5 (12.2 to 13.0) | 15.2 (14.8 to 15.6) | 16.7 (16.3 to 17.2) | 16.8 (16.4 to 17.2) |
|  | Second Least | 18.2 (17.6 to 18.8) | 17.6 (17.1 to 18.1) | 17.0 (16.5 to 17.5) | 16.8 (16.3 to 17.3) | 17.3 (16.9 to 17.8) | 12.4 (12.0 to 12.8) | 15.7 (15.3 to 16.1) | 16.8 (16.4 to 17.3) | 17.4 (16.9 to 17.8) |
|  | Neither | 18.5 (18.0 to 19.0) | 18.0 (17.5 to 18.5) | 16.6 (16.1 to 17.0) | 17.2 (16.8 to 17.7) | 17.5 (17.0 to 17.9) | 12.8 (12.5 to 13.2) | 15.8 (15.4 to 16.2) | 17.0 (16.7 to 17.4) | 17.6 (17.3 to 18.0) |
|  | Second Most | 18.1 (17.7 to 18.5) | 17.3 (16.9 to 17.7) | 16.9 (16.5 to 17.3) | 17.4 (17.0 to 17.8) | 17.6 (17.2 to 18.0) | 13.3 (13.0 to 13.6) | 16.0 (15.7 to 16.3) | 17.3 (17.0 to 17.7) | 17.9 (17.6 to 18.3) |
|  | Most | 17.7 (17.3 to 18.1) | 17.7 (17.3 to 18.1) | 17.1 (16.7 to 17.5) | 17.5 (17.1 to 17.8) | 17.4 (17.1 to 17.8) | 13.0 (12.7 to 13.3) | 15.7 (15.4 to 16.1) | 17.3 (17.0 to 17.7) | 17.9 (17.6 to 18.3) |
| White | Least | 26.8 (26.7 to 26.9) | 26.2 (26.1 to 26.3) | 25.2 (25.2 to 25.3) | 25.7 (25.6 to 25.8) | 26.2 (26.1 to 26.3) | 19.6 (19.5 to 19.7) | 23.2 (23.1 to 23.2) | 25.1 (25.1 to 25.2) | 26.0 (25.9 to 26.1) |
|  | Second Least | 26.6 (26.5 to 26.7) | 25.9 (25.8 to 25.9) | 24.8 (24.7 to 24.9) | 25.2 (25.1 to 25.2) | 25.8 (25.7 to 25.9) | 19.3 (19.3 to 19.4) | 22.8 (22.7 to 22.8) | 24.7 (24.6 to 24.8) | 25.6 (25.5 to 25.7) |
|  | Neither | 26.1 (26.0 to 26.1) | 25.4 (25.3 to 25.5) | 24.1 (24.0 to 24.2) | 24.5 (24.4 to 24.6) | 25.1 (25.0 to 25.1) | 18.8 (18.8 to 18.9) | 22.2 (22.2 to 22.3) | 24.1 (24.0 to 24.2) | 25.0 (24.9 to 25.1) |
|  | Second Most | 25.7 (25.6 to 25.8) | 25.1 (25.0 to 25.2) | 24.0 (24.0 to 24.1) | 24.3 (24.2 to 24.4) | 24.7 (24.6 to 24.7) | 18.6 (18.5 to 18.7) | 22.0 (22.0 to 22.1) | 23.7 (23.6 to 23.7) | 24.5 (24.4 to 24.6) |
|  | Most | 26.0 (25.9 to 26.1) | 25.6 (25.5 to 25.7) | 24.5 (24.5 to 24.6) | 24.7 (24.6 to 24.8) | 24.9 (24.8 to 25.0) | 18.8 (18.8 to 18.9) | 22.5 (22.4 to 22.6) | 23.9 (23.8 to 24.0) | 24.7 (24.7 to 24.8) |
| English Region | London | 24.1 (24.0 to 24.1) | 23.6 (23.5 to 23.6) | 22.8 (22.7 to 22.9) | 23.2 (23.1 to 23.2) | 23.2 (23.1 to 23.2) | 16.8 (16.8 to 16.9) | 20.0 (19.9 to 20.0) | 21.7 (21.7 to 21.8) | 22.4 (22.3 to 22.4) |
|  | Midlands/East | 23.9 (23.8 to 23.9) | 23.3 (23.2 to 23.3) | 22.6 (22.5 to 22.6) | 23.2 (23.2 to 23.3) | 23.9 (23.8 to 24.0) | 17.8 (17.7 to 17.8) | 21.3 (21.3 to 21.4) | 23.2 (23.1 to 23.2) | 24.4 (24.3 to 24.4) |
|  | North England | 25.0 (24.9 to 25.1) | 24.5 (24.4 to 24.6) | 23.7 (23.6 to 23.7) | 23.9 (23.8 to 23.9) | 24.2 (24.2 to 24.3) | 18.3 (18.2 to 18.3) | 21.9 (21.9 to 22.0) | 23.4 (23.3 to 23.4) | 24.1 (24.1 to 24.2) |
|  | South England | 25.1 (25.0 to 25.2) | 24.6 (24.6 to 24.7) | 23.3 (23.3 to 23.4) | 23.4 (23.3 to 23.5) | 23.8 (23.7 to 23.8) | 18.0 (17.9 to 18.0) | 21.0 (21.0 to 21.1) | 22.7 (22.6 to 22.7) | 23.5 (23.4 to 23.5) |

Table S4	Mean Variation in A&G and Direct Referrals by Region: 2023

| Region | Mean difference between A&G and direct referral rate | Range | Correlation* |
| --- | --- | --- | --- |
| Total | 22.6% | 5.8% to 34.3% | 0.010 (0.925) |
| London/South England | 21.3% | 17.6% to 28.7% | -0.274 (0.243) |
| Midlands/East England | 23.5% | 12.6% to 29.4% | 0.052 (0.812) |
| North England | 22.7% | 5.8% to 34.3% | 0.062 (0.691) |

* Pearson’s product moment correlation coefficient

Table S5	Characteristics of those with/without Direct Referrals within ±12 months of A&G

|  |  |  | Direct Referral | | Before A&G only | | After A&G only | |
| --- | --- | --- | --- | --- | --- | --- | --- | --- |
| A&G with Direct Referral ±12 months | | A&G Only | Before A&G only | After A&G only | RRR (95% CI) | aRRR (95% CI) | RRR (95% CI) | aRRR (95% CI) |
| Total, n (%) | | 34,496 (8) | 305,231 (70) | 94,902 (22) | --- | --- | --- | --- |
| Gender, n (%) | Females | 19,173 (7) | 181,385 (71) | 56,309 (22) | [reference] | [reference] | [reference] | [reference] |
|  | Males | 15,323 (9) | 123,846 (70) | 38,593 (22) | 0.85 (0.83, 0.88) | 0.85 (0.83, 0.88) | 0.86 (0.83, 0.88) | 0.85 (0.83, 0.88) |
| Age Bands, n (%) | 0-4 years | 1,214 (14) | 5,812 (66) | 1,779 (20) | 0.56 (0.49, 0.63) | 0.58 (0.52, 0.66) | 0.55 (0.49, 0.62) | 0.57 (0.51, 0.64) |
|  | 5-9 years | 2,004 (13) | 9,948 (66) | 3,067 (20) | 0.58 (0.52, 0.64) | 0.60 (0.54, 0.66) | 0.58 (0.53, 0.63) | 0.59 (0.54, 0.64) |
|  | 10-14 years | 1,900 (13) | 10,007 (67) | 3,014 (20) | 0.61 (0.56, 0.67) | 0.63 (0.57, 0.69) | 0.60 (0.55, 0.65) | 0.61 (0.56, 0.67) |
|  | 15-24 years | 2,814 (10) | 18,113 (68) | 5,879 (22) | 0.75 (0.69, 0.82) | 0.75 (0.69, 0.82) | 0.79 (0.73, 0.84) | 0.80 (0.75, 0.86) |
|  | 25-34 years | 3,359 (9) | 26,447 (70) | 8,099 (21) | 0.92 (0.87, 0.97) | 0.92 (0.87, 0.97) | 0.91 (0.85, 0.96) | 0.92 (0.87, 0.98) |
|  | 35-44 years | 4,156 (8) | 35,694 (70) | 11,051 (22) | [reference] | [reference] | [reference] | [reference] |
|  | 45-54 years | 5,025 (8) | 45,522 (70) | 14,285 (22) | 1.05 (1.00, 1.11) | 1.07 (1.02, 1.12) | 1.07 (1.02, 1.12) | 1.10 (1.05, 1.16) |
|  | 55-64 years | 5,119 (7) | 49,646 (71) | 15,579 (22) | 1.13 (1.06, 1.20) | 1.17 (1.10, 1.24) | 1.14 (1.08, 1.21) | 1.22 (1.15, 1.29) |
|  | 65-74 years | 4,747 (7) | 48,261 (71) | 15,364 (22) | 1.18 (1.10, 1.28) | 1.25 (1.16, 1.35) | 1.22 (1.13, 1.31) | 1.32 (1.23, 1.42) |
|  | 75-84 years | 3,067 (5) | 40,712 (73) | 12,330 (22) | 1.55 (1.40, 1.71) | 1.64 (1.50, 1.79) | 1.51 (1.39, 1.65) | 1.65 (1.52, 1.80) |
|  | 85+ years | 1,091 (5) | 15,069 (73) | 4,455 (22) | 1.61 (1.44, 1.80) | 1.70 (1.53, 1.89) | 1.54 (1.38, 1.71) | 1.67 (1.50, 1.87) |
| Ethnicity, n (%) | Asian | 2,730 (7) | 24,419 (64) | 11,022 (29) | 1.02 (0.80, 1.30) | 1.10 (0.87, 1.39) | 1.55 (1.28, 1.88) | 1.74 (1.45, 2.10) |
|  | Black | 1,142 (6) | 12,854 (71) | 4,134 (23) | 1.28 (0.96, 1.71) | 1.30 (1.00, 1.70) | 1.39 (1.13, 1.71) | 1.55 (1.27, 1.89) |
|  | Mixed/multiple | 533 (8) | 4,443 (68) | 1,531 (24) | 0.95 (0.80, 1.13) | 1.18 (0.99, 1.40) | 1.10 (0.96, 1.27) | 1.44 (1.25, 1.66) |
|  | White | 29,472 (8) | 258,742 (71) | 76,769 (21) | [reference] | [reference] | [reference] | [reference] |
|  | Other | 73 (7) | 673 (67) | 257 (26) | 1.05 (0.78, 1.41) | 1.25 (0.93, 1.67) | 1.35 (1.01, 1.81) | 1.70 (1.27, 2.27) |
|  | Unknown | 546 (9) | 4,100 (70) | 1,189 (20) | 0.86 (0.68, 1.07) | 0.97 (0.78, 1.21) | 0.84 (0.63, 1.10) | 0.95 (0.73, 1.25) |
| IMD quintile, n (%) | Least | 6,836 (7) | 63,806 (70) | 20,588 (23) | [reference] | [reference] | [reference] | [reference] |
|  | Second least | 7,956 (9) | 63,520 (69) | 20,898 (23) | 0.86 (0.70, 1.04) | 0.87 (0.72, 1.06) | 0.87 (0.76, 1.00) | 0.88 (0.77, 1.01) |
|  | Mid | 7,383 (8) | 62,252 (69) | 20,663 (23) | 0.90 (0.72, 1.13) | 0.92 (0.74, 1.16) | 0.93 (0.79, 1.10) | 0.92 (0.79, 1.08) |
|  | Second most | 6,780 (8) | 62,207 (70) | 19,614 (22) | 0.98 (0.73, 1.32) | 1.01 (0.75, 1.36) | 0.96 (0.77, 1.20) | 0.94 (0.76, 1.16) |
|  | Most | 5,500 (8) | 53,326 (74) | 13,071 (18) | 1.04 (0.77, 1.40) | 1.09 (0.80, 1.47) | 0.79 (0.64, 0.97) | 0.79 (0.64, 0.97) |
|  | Missing | 41 (18) | 120 (52) | 68 (30) | 0.31 (0.15, 0.65) | 0.31 (0.14, 0.70) | 0.55 (0.34, 0.89) | 0.57 (0.33, 0.97) |
| Year, n (%) * | 2015 | 1,001 (10) | 6,360 (66) | 2,243 (23) | 0.67 (0.45, 0.98) | 0.69 (0.47, 1.00) | 0.87 (0.61, 1.25) | 0.92 (0.65, 1.31) |
|  | 2016 | 1,202 (10) | 7,770 (66) | 2,833 (24) | 0.68 (0.49, 0.94) | 0.68 (0.50, 0.94) | 0.92 (0.65, 1.30) | 0.94 (0.67, 1.32) |
|  | 2017 | 1,758 (11) | 10,525 (65) | 3,862 (24) | 0.63 (0.44, 0.91) | 0.63 (0.44, 0.91) | 0.86 (0.60, 1.22) | 0.88 (0.62, 1.24) |
|  | 2018 | 2,350 (7) | 23,375 (73) | 6,233 (20) | 1.05 (0.76, 1.45) | 1.04 (0.75, 1.43) | 1.03 (0.81, 1.33) | 1.05 (0.82, 1.34) |
|  | 2019 | 3,367 (8) | 27,537 (68) | 9,712 (24) | 0.86 (0.69, 1.07) | 0.85 (0.68, 1.05) | 1.13 (0.95, 1.33) | 1.13 (0.96, 1.33) |
|  | 2020 | 6,095 (7) | 58,136 (69) | 19,758 (24) | 1.00 (0.87, 1.16) | 1.00 (0.86, 1.16) | 1.26 (1.14, 1.40) | 1.26 (1.14, 1.40) |
|  | 2021 | 8,984 (8) | 78,850 (70) | 25,299 (22) | 0.92 (0.85, 1.00) | 0.93 (0.85, 1.01) | 1.10 (1.03, 1.17) | 1.10 (1.03, 1.17) |
|  | 2022 | 9,739 (8) | 92,678 (73) | 24,962 (20) | [reference] | [reference] | [reference] | [reference] |

* 2022 omitted due to low numbers with >12 months follow-up.
Abbreviations: RR = risk ratio; aRR = adjusted risk. * Before A&G defined as direct referral recorded on -122 to 0 days; after A&G defined as direct referral recorded on 1-122 days.

Table S6	Comparison of Direct Referral Rates from CPRD Aurum to e-RS Dashboard

| Year | ONS England population, n | e-RS referrals, n | e-RS referral rate  per 100 persons | CPRD Aurum referral rate per 100 persons |
| --- | --- | --- | --- | --- |
| 2020 | 56,326,000 | 9,512,554 | 16.9 | 17.8 |
| 2021 | 56,554,900 | 12,720,787 | 22.5 | 21.1 |
| 2022 | 57,112,500 | 14,115,174 | 24.7 | 22.8 |
| 2023 | 57,690,300 | 14,918,959 | 25.9 | 23.6 |

ONS = Office for National Statistics; e-RS = e-Referral System; CPRD = Clinical Practice Research Datalink.
